# Predictive Modelling to Differentiate Bacterial and Viral cases of Childhood Pneumonia in Kilifi, Kenya using Protein Markers and Clinical Data

**DOI:** 10.64898/2026.04.08.26350312

**Authors:** Cynthia Matuli, Jacqueline M. Waeni, Elijah T. Gicheru, Charles J. Sande, Katherine Gallagher

## Abstract

**Background:** To date, accessible diagnostic tools to identify whether a patient’s pneumonia is a bacterial, or viral infection, are not accurate or timely enough to prevent preemptive antibiotic administration. Relying on single biomarkers or clinical presentations has been insufficient. We aimed to incorporate a wide range of novel biomarkers and clinical presentations in a multivariable model and validate its capacity to differentiate cases of bacterial and viral pneumonia.

**Methods:** Data from 457 children aged 2–59 months, admitted to Kilifi County Referral Hospital, Kenya, with bacterial (n = 229) and viral (n = 228) infections, were used to develop and validate a predictive multivariable Poisson regression model to differentiate pneumonia etiology. The Receiver Operating Characteristic curve was used to assess biomarker performance and validate the model internally.

**Results:** Sixty-three percent (63%) of the children presented with severe pneumonia. 72% with viral pneumonia had severe pneumonia, compared to 54% with bacterial pneumonia who had severe pneumonia. In crude analyses, chest-wall indrawing, cough, convulsions, crackles, angiotensinogen, and Serpin Family A Member 1 were significantly associated with pneumonia etiology, controlling for age. However, only chest-wall indrawing remained significant in multivariable analyses after controlling for age. The model demonstrated fair, but inadequate, discrimination, with an Area Under the Curve of 0.61.

**Conclusion:** Among the children admitted to hospital with WHO defined pneumonia, a wide range of biomarkers and clinical presentations still failed to distinguish bacterial from viral pneumonia.

## Introduction

Globally, the incidence of pneumonia in the pediatric population has significantly decreased following the widespread administration of pneumococcal conjugate vaccines (PCVs) and *Haemophilus influenzae* type b (Hib) vaccines [1,2]. However, pneumonia remains one of the leading causes of pediatric morbidity and mortality, particularly in sub-Saharan Africa. As bacterial infections have decreased, severe viral infections now form a larger proportion of the case load presenting to hospital [1,3]. Guidelines still recommend the empirical use of antibiotics, as this has been instrumental in preventing bacterial infection-related deaths; however, it is thought unnecessary use of antibiotics for viral infections has inadvertently fueled antimicrobial resistance (AMR) [4,5]. Diagnostic tools to guide the rational use of antibiotics for pneumonia treatment are needed.

The distinction between bacterial and viral pneumonia remains challenging, especially in Low-and Middle-Income countries (LMICs). An accurate distinction between bacterial and viral pathogens is crucial for guiding antibiotic treatment [5]. However, conventional methods to identify bacterial infection, such as blood and sputum cultures, are invasive, have limited sensitivity, and are time-consuming [6,7]. Meanwhile, molecular testing, such as Polymerase Chain Reaction (PCR) [6,7], provides higher sensitivities and specificities, but is expensive, and only detects specific pathogens. Transcriptomics has been used to accurately distinguish bacterial from viral pneumonia with high sensitivity and specificity [8,9]; however, it remains impractical in LMICs due to its high cost and the need for advanced laboratory infrastructure [9].

Scalable and more affordable methods to distinguish bacterial pneumonia from viral pneumonia remain elusive [10,11]. Individuals with bacterial pneumonia and those with viral pneumonia present with overlapping clinical signs and symptoms and biomarker levels making the use of individual symptoms or concentrations of solitary markers suboptimal [12–14]. Concentrations of biomarkers such as C-reactive protein (CRP) and procalcitonin (PCT), have in the past been significantly higher in bacterial pneumonia compared to viral pneumonia cases [15]. However, these biomarkers had low sensitivities (CRP = 0.70, PCT = 0.66). CRP lacks etiological specificity and is elevated by co-infections such as malaria [6,10,12], obesity, and cardiovascular diseases [16]. PCT can be falsely low in localized infections, such as empyema, due to the time lag between infection onset and its peak, and is also elevated by coinfections, surgery, or trauma [11,17]. White blood cell counts (WBC), absolute neutrophil count (ANC), and monocyte values vary by ethnicity, with individuals of African ancestry having significantly lower counts, which limits generalizability [18].

Combining a wide range of biomarkers with clinical presentations may result in better diagnostic accuracy [5,19,20]. Very few studies have attempted this, especially in sub-Saharan Africa and in the pediatric population and the resultant models have had limited predictive capacity, have included only a few serological biomarkers and have had small study sample sizes [24]. This study integrates a wide range of novel protein biomarkers, demographic, anthropometric, and clinical data into a single model to predict whether a child presenting with pneumonia is likely to have a bacterial or viral infection.

## METHODS

### Study setting and population

Data on children presenting to Kilifi County Referral Hospital (KCRH) with pneumonia were derived from the long-term surveillance of RSV in childhood lower respiratory tract infection study, conducted between January 2007 and August 2020. This project sampled children less than 5 years of age who were either febrile or presented with sepsis or pneumonia for biomarker discovery and validation [8]. Children were excluded if born prematurely, were less than 1 week of age, or had a history of congenital heart disease. For the analysis presented here, this nested case-control study selected data from children aged 2-59 months who met the 2013 WHO definition of pneumonia [18].

### Study procedures

Under the surveillance protocol, anthropometric (e.g., weight, height), clinical (e.g., cough, cyanosis, hypoxemia), and demographic data (e.g., age, gender), blood samples, and nasopharyngeal/oropharyngeal (NP/OP) swabs were collected at admission. NP/OP swabs were used to test for 15 common childhood viral infections, including adenovirus, parainfluenza virus (1,2,3, and 4), RSV (A and B), influenza (A, B, and C), rhinovirus, human metapneumovirus, and coronaviruses (OC43, NL63, and E229) using multiplex real-time PCR. Blood specimens were cultured for bacteria, and a blood slide was prepared to determine malaria infection status. Blood samples were analyzed with a complete blood count (WBC, red blood cell counts (RBC), platelets, neutrophils, eosinophils, and basophils). The biomarker discovery study tested the blood samples from a subset of children and quantified 418 plasma proteins using high-performance liquid chromatography tandem mass spectrometry (HPLC-MS/MS) [11]. CRP and 6 other plasma proteins appeared predictive of infectious aetiology: angiotensinogen (AGT), Serpin Family A Member 1 (SERPINA1), Serpin Family A Member 3 (SERPINA3), Paraoxonase-1 (PON1), Histidine Rich Glycoprotein (HRG/B2R8I2), and lipopolysaccharide-binding protein (LBP) [11] and therefore considered for etiology differentiation for this study. Samples from the whole cohort of children were then tested for CRP, PCT, and these 6 proteins using the same mass spectrometry. The ILab Aries clinical biochemistry analyzer was used to measure a subset of 100 CRP samples which were converted to concentrations [8]. Laboratory sample testing was conducted at the KEMRI-Wellcome Trust Research Programme (KWTRP) in Kilifi, Kenya.

For the secondary data analysis presented in this paper, pneumonia cases were classified as viral, bacterial, bacterial-viral co-infections, and unknown infections (Figure 1). For clinical relevance, we combined children with evidence of bacterial infections, bacterial-viral co-infections, and unknown infections into a single group (possible bacterial infections) that would likely need to receive empirical antibiotic therapy if presenting to a healthcare facility. Children with evidence of viral infections known to cause pneumonia (adenovirus, parainfluenza virus (1,2,3, and 4), RSV (A and B), influenza (A, B, and C), and human metapneumovirus) were defined as the probable viral pneumonia group.

**Figure 1:**
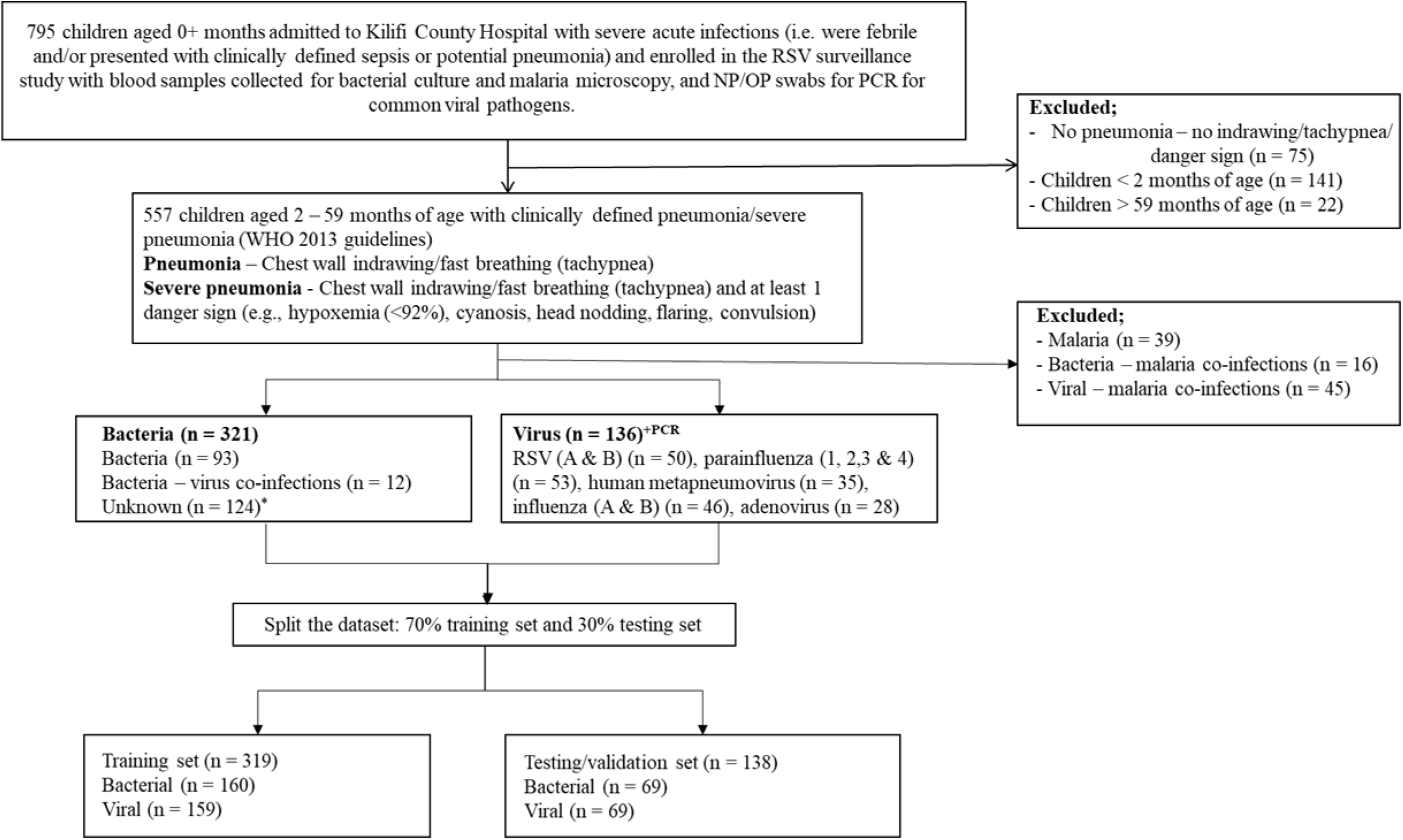
Study participant flowchart. *The unknowns include individuals PCR positive for Rhinovirus, and endemic hCoV strains (OC43, NL63, and 229E) not thought to cause pneumonia, and culture positive for known contaminants e.g., Streptococcus hominis, and/or those which were negative on PCR targets and negative on blood culture. +PCRPositive PCR test for RSV A and B, Parainfluenza 1,2,3,4, Adenovirus, Influenza A, B, C, or Human metapneumovirus (hMPV).

### Statistical analyses

Variables such as cough duration were missing in >25% of participants and excluded from the analysis. Using the full dataset, the Receiver Operating Characteristic (ROC) curve was used to identify optimal cutoff points for the mean fluorescent intensity (MFI) values of biomarkers, enabling categorization into categorical variables with the highest sensitivity, specificity, and overall discriminatory ability. The cutpointr package in R was utilized to determine the optimal threshold of each biomarker using the Youden index (sensitivity + specificity – 1). To enhance the robustness of the estimates, the optimal threshold was derived using 1000 bootstrap resamples. Cut-off points for biomarkers were also identified when the dataset was stratified by age (<1 and ≥1 years, <2 and ≥2 years, and <3 and ≥3 years) to determine if cut-offs varied by age. The Variance Inflation Factor (VIF) was used to evaluate multicollinearity among variables. Highly correlated variables (VIF >2) were assessed for their discriminatory ability. The variable with the lower AUC was dropped.

### Model Development

The dataset was randomly split into two datasets, stratified by etiology. 70% of the data was used for training and 30% for model validation. Age was considered *a priori* as a confounder in every stage of the analysis. A modified Poisson regression model was used to assess the association between every predictor and the outcome. Covariates with a likelihood ratio test (LRT) p-value < 0.2 in the crude analyses were considered to improve model fit and were therefore included in selection for the multivariable predictive model. The multivariable Poisson regression model was fitted using backward selection. Variables were iteratively removed from the multivariate model, and variables that significantly improved the model’s fit (LRT p-value < 0.05) were retained in the final model. The overall sensitivity, specificity, and discriminatory power (AUC) of the final model in distinguishing between bacterial and viral pneumonia cases were assessed.

### Model Validation

The model’s ability to distinguish between bacterial and viral pneumonia cases was assessed using Area Under the Curve (AUC) values on the validation dataset. Other published predictive models were also run on the validation dataset to determine comparative discriminatory power. An AUC of 0.5 indicated that the model cannot discriminate beyond chance; a value greater than 0.7 was considered moderate discrimination, while a value greater than 0.80 was regarded as good discrimination [22]. The model’s calibration was assessed using the calibration intercept which measures how close the predicted risks are to the observed event (underestimation or overestimation) and the calibration slope which determines whether there’s potential model overfit or underfit [21]. We externally validated a previously published model [15], developed in a study conducted in Indonesian children 2-59 months of age with pneumonia (WHO 2014 definition) and hospitalized within 24 hours of admission, by applying their logistic regression model to our dataset without any modification. We defined the variables in their final model as defined in their publication without refitting or recalibrating their model. Another study [9] conducted in Western Australian children ≤17 years of age, recommended a CRP cut off of 72mg/L with the presence of fever (≥ 38° C) or absence of rhinorrhea. We validated these models using the CRP data that was analysed on the ILab Aries clinical biochemistry analyzer. All statistical analyses were conducted using R software version 4.5.1, and variable significance was evaluated at a 5% significance level.

### Ethics approval

The long-term surveillance of RSV in childhood lower respiratory tract infections study (SSC 1055) was approved by the Kenya Medical Research Institute Scientific Ethics Research Unit (KEMRI–SERU) and conducted in accordance with the principles of Good Clinical Practice and Good Clinical Laboratory Practice (GCLP). Written informed consent was obtained from the participant’s respective legal guardian or next of kin, and the study adhered to the principles outlined in the Declaration of Helsinki. For this nested study, data were shared with the approval of the study Principal Investigator and the KWTRP data governance committee.

## RESULTS

Among the 457 children enrolled in this study, 228 (49.9%) had a viral infection, 93 (20.4%) had a bacterial infection, 12 (2.6%) had a bacterial-viral co-infection, and 124 (27.1%) had an unknown etiology. Among the 228 with a viral infection, the most identified viral pathogens were RSV-A (n = 39, 17.1%), parainfluenza-3 (n = 36, 15.8%), human metapneumovirus (n = 35, 15.4%), influenza-A (n = 34, 14.9%), and adenovirus (n = 28, 12.3%). The most identified bacterial pathogens from blood cultures were *Escherichia coli* (n = 17, 7.4%)*, Streptococcus pneumoniae* (n = 17, 7.4%), *Staphylococcus aureus* (n = 11, 4.8%), and *Haemophilus influenza* (n = 8, 3.5%). (Supplementary figures 1 and 2).

### Demographic, laboratory, and clinical characteristics

More than half (63%) of the study population had severe pneumonia compared to non-severe pneumonia (37%). A higher proportion of children with probable viral pneumonia presented with severe pneumonia (72%), compared to children with possible bacterial pneumonia (54%). Cell counts of WBC, neutrophils, and the median fluorescent intensity (MFI) units of AGT, LBP, PCT, and SERPINA1 were higher in the bacterial group, while the cell counts of RBC, hemoglobin concentrations, and MFI units of SERPINA3 were higher in the viral group.

### Predictive capacity of each biomarker to differentiate bacterial from viral infections

Based on this nested study’s case definition, no biomarker could differentiate between bacterial and viral infections beyond random chance (Table 1). Figure 2 shows the high degree of overlap observed across the bacterial and viral biomarker levels (Figure 2).

**Figure 2:**
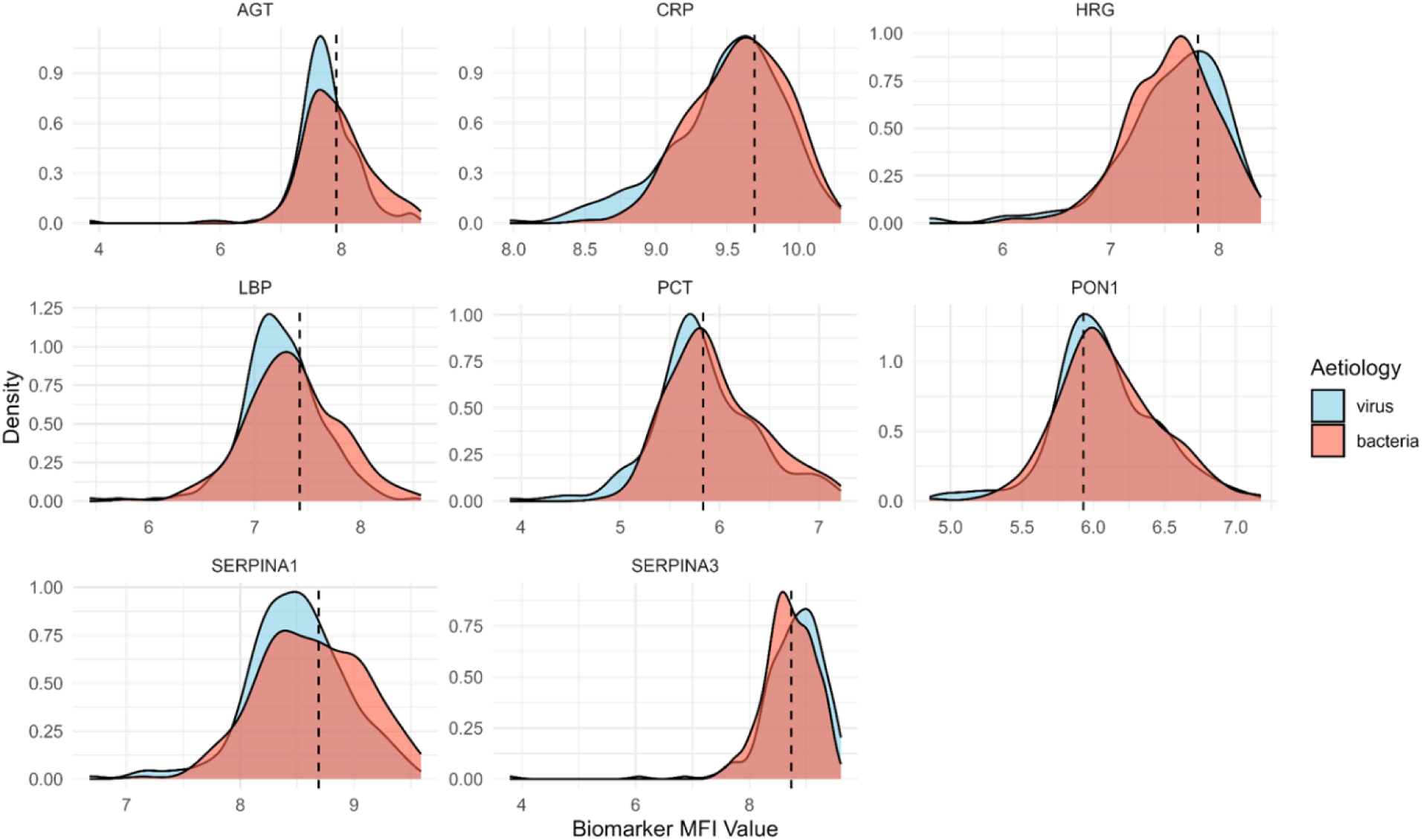
The distribution of median flourescent intensity units among all possible bacterial infections and probable viral infections Overlap of biomarker MFI levels in bacterial and viral infections. The dotted lines indicate the biomarkers optimal thresholds (cutoff values).

**Table 1:** Biomarker thresholds derived when comparing all possible bacterial cases and probable viral cases, and their sensitivity and specificities in differentiating between all possible bacterial cases and probable viral cases.

| Biomarker | Threshold (MFI) | Sensitivity | Specificity | PPV | NPV | AUC [95% CI] |
| --- | --- | --- | --- | --- | --- | --- |
| AGT | 7.9144 | 0.46 | 0.72 | 0.62 | 0.57 | 0.58 [0.53, 0.64] |
| CRP | 9.2097 | 0.87 | 0.22 | 0.53 | 0.63 | 0.55 [0.50, 0.60] |
| PCT | 5.8311 | 0.58 | 0.57 | 0.58 | 0.58 | 0.56 [0.51, 0.62] |
| LBP | 7.4229 | 0.42 | 0.71 | 0.60 | 0.55 | 0.58 [0.52, 0.63] |
| HRG | 7.7156 | 0.63 | 0.46 | 0.54 | 0.56 | 0.53 [0.48, 0.58] |
| PON1 | 6.1453 | 0.42 | 0.65 | 0.55 | 0.53 | 0.52 [0.47, 0.58] |
| SERPINA1 | 8.6896 | 0.46 | 0.71 | 0.61 | 0.56 | 0.58 [0.53, 0.64] |
| SERPINA3 | 8.7402 | 0.54 | 0.61 | 0.58 | 0.57 | 0.57 [0.51, 0.62] |
|  | Threshold (counts) | Sensitivity | Specificity | PPV | NPV | AUC [95% CI] |
| WBC | 17.65 | 0.45 | 0.74 | 0.64 | 0.57 | 0.58 [0.53, 0.64] |
| Neutrophils | 8.715 | 0.43 | 0.78 | 0.64 | 0.61 | 0.57 [0.51, 0.64] |
| Lymphocytes | 6.695 | 0.78 | 0.31 | 0.49 | 0.62 | 0.54 [0.48, 0.60] |

Area Under the Curve (AUC) represents the discriminatory power of each predictor between the bacterial and viral cases of pneumonia. Sensitivity is the proportion of true bacterial cases accurately identified by each predictor. Specificity is the proportion of true viral cases accurately identified by each predictor. Positive Predictive Value (PPV) is the proportion of cases predicted as bacterial that are truly bacterial. Negative Predictive Value (NPV) is the proportion of cases predicted as viral that are truly not bacterial.

### Modified Poisson regression analyses

In crude analyses controlling for age, restricted to the training set, mid-upper arm circumference, chest-wall indrawing, cough, convulsion, crackles, AGT (in continuous and categorical forms), and SERPINA1 (in a categorical form that used different cut-offs for children <1 and ≥1 year of age) were predictive of pneumonia etiology (Table 2). In multivariable analyses controlling for age, only lower chest-wall indrawing (LCWI) (aRR = 0.75, 95% CI [0.61, 0.93], p-value = 0.0081) remained a significant predictor, with a higher prevalence in the viral pneumonia group. After controlling for age and LCWI, none of the other clinical signs or symptoms, nor biomarker levels were predictive of etiology.

**Table 2:**
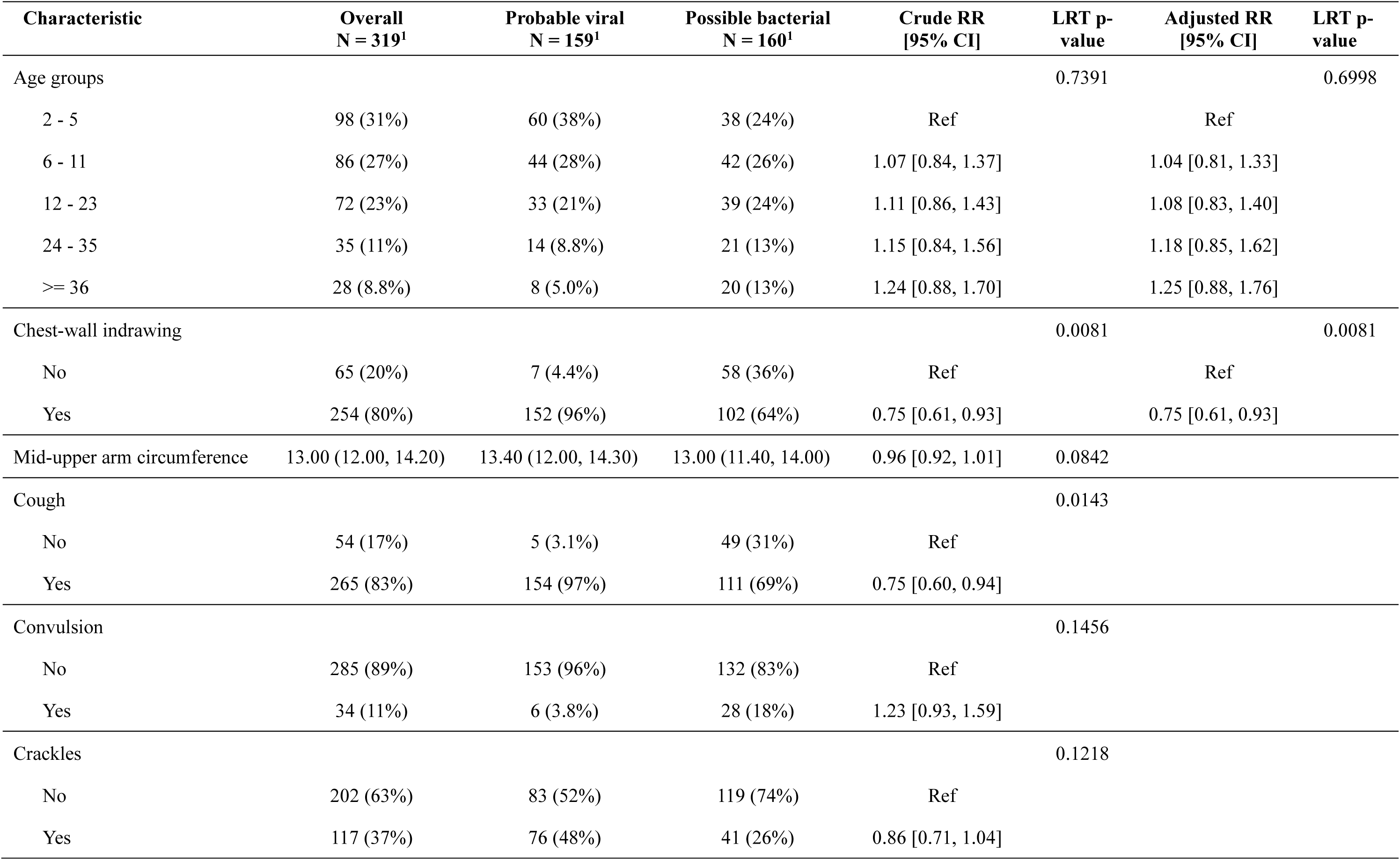
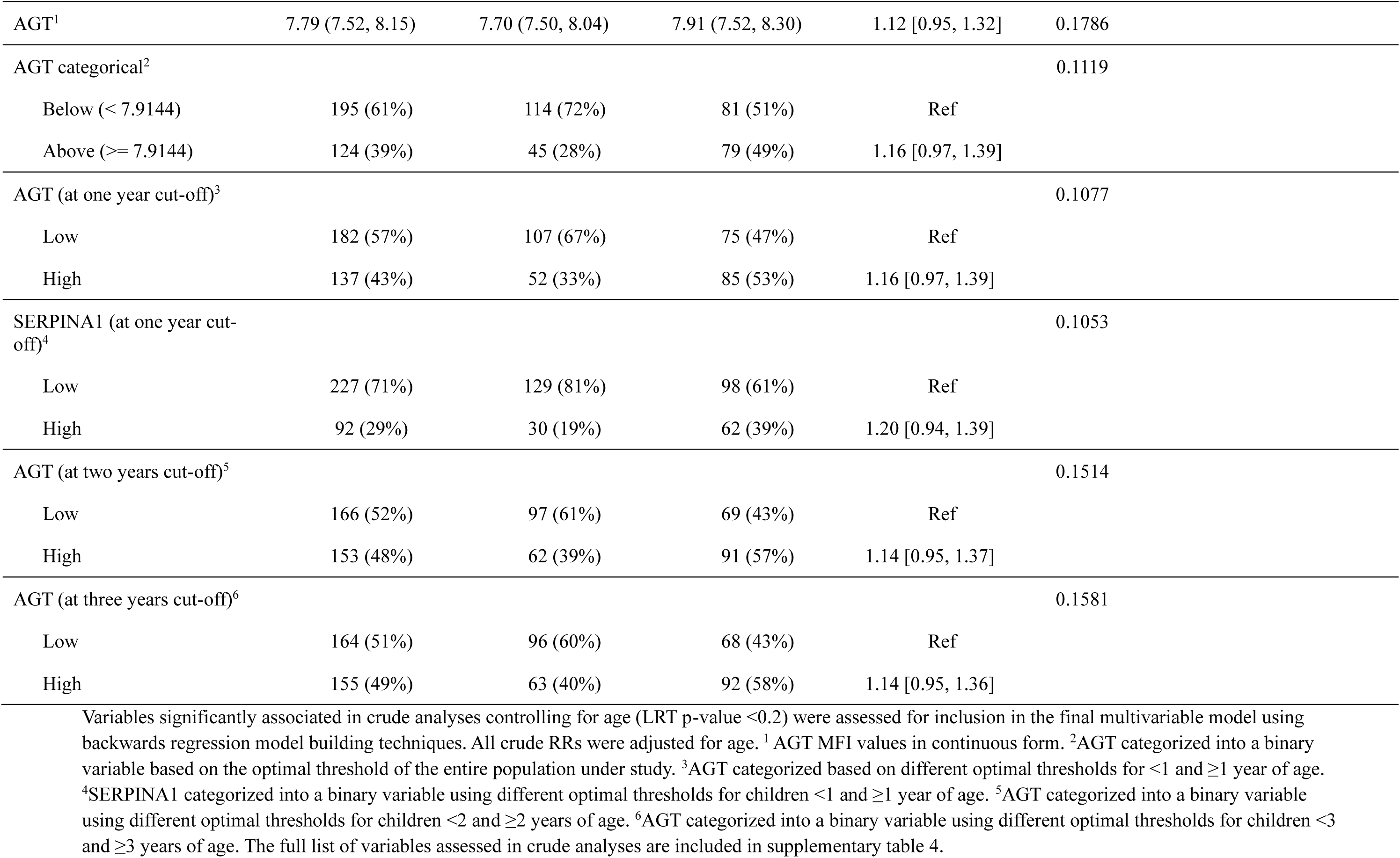
Predictors of pneumonia etiology identified in the training dataset using a multivariable Poisson regression model.

The multivariable model including age and LCWI exhibited moderate discrimination (AUC = 0.72, 95% CI [0.66, 0.77], with a sensitivity of 0.55 and a specificity of 0.84) on the training dataset. The model performed poorly on the validation dataset (AUC = 0.61, 95% CI [0.52, 0.70]; with a sensitivity of 0.50 and a specificity of 0.68) (Figure 3).

**Figure 3:**
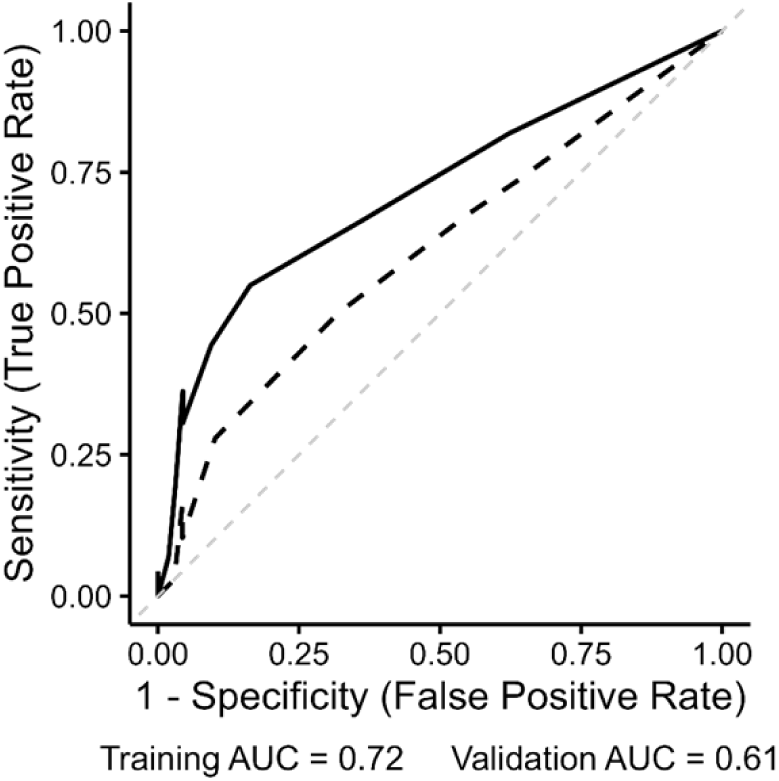
The predictive capacity of the final multivariable model displayed as the Receiver Operating Curve (ROC) when applied to the training dataset (solid line, AUC 0.72) and the validation dataset (dotted line, AUC 0.61)

### Comparison with other available models

A study conducted in Indonesia [15] by Farida et al., identified CRP (≥ 5.70mg/L), presence of fever (≥ 38°C), and rainy season as predictors of pneumonia etiology. The combination of these three factors exhibited poor predictive capacity and could not discriminate beyond random chance in our validation study (AUC of 0.59 (95% CI [0.46, 0.71])). Bhuiyan et al.’s study [9] identified CRP (≥ 72mg/L), fever (≥ 38°C) and presence or absence of rhinorrhea as significant predictors of etiology. When these factors were combined, the model exhibited moderate predictive capacity in our validation dataset (AUC = 0.74, (95% CI [0.64, 0.85] with a sensitivity of 0.55 and a specificity of 0.86) (Supplementary Table 6).

## DISCUSSION

In this nested case-control study, we utilized a dataset including data on a wide range of clinical signs and symptoms, cell counts, and novel biomarker levels to develop a multivariable predictive model and evaluated its ability to discriminate between bacterial and viral pneumonia in children presenting with WHO defined pneumonia. The model incorporated eight (8) biomarkers, some of which have not been assessed for this purpose before, to evaluate their incremental value in etiology prediction and account for any underlying biological pathways not captured by conventional biomarkers. 63% of the study participants were severely ill, reflecting the severe case mix presenting to hospital in Kilifi, and more broadly, sub-Saharan Africa [22,23], morbidity is underpinned by poverty, malnutrition, and inaccessibility to healthcare facilities.

In crude analyses, higher MFI units of AGT and SERPINA1 correlated with increased probability of an individual having bacterial pneumonia. However, neither of the biomarkers remained significantly predictive of bacterial infections after controlling for age and chest-wall indrawing. In internal validation, the final model including age and LCWI did not have the predictive capacity to guide clinical decision-making (AUC = 0.61, 95% CI [0.52, 0.70], sensitivity = 0.50, specificity = 0.68).

Other published models developed for the same purpose have reported equally poor performance on internal validation. Performance of these models did not improve when we externally validated them on our dataset. The study by Farida et al., [15] was conducted in a population with similar age-group and case mix to our population. However, their outcome of interest was defined differently to ours as they excluded unknown infections. Despite this, their model performed equally badly on our population as in their own internal validation. The model identified by Bhuiyan et al. [9] also performed poorly on our population.

Differentiating bacterial from viral pneumonia remains a challenge. Incorporating novel biomarker levels in our study did not improve the predictive capacity compared to other models which included simply clinical and demographic data. The distribution of MFI units significantly overlapped in bacterial and viral cases. To explore possible reasons why, after controlling for age and LCWI, biomarker levels were not predictive, we assessed interactions between biomarkers predictive capacity and pneumonia severity and interactions with age. CRP, AGT, HRG, and SERPINA1 had significant interactions with pneumonia severity; higher values were more predictive of pneumonia etiology in non-severe cases than in severe cases (supplementary table 8) as also reported by Wu et al. [25]. Other studies have identified that levels of SERPINA1, PON1, and LBP are higher in bacterial infections and do identify patients with CAP from other patients [7,30–34], however, these markers do not adequately differentiate between bacterial and viral etiologies. HRG has been shown to exhibit an antifungal role and levels negatively correlate with disease severity [35]. Previous studies have reported interactions between CRP and LBP and age [16,27–29], we assessed each biomarker using different cut-offs for different ages; however, this did not significantly improve their predictive capacity.

This analysis, as with others before it, has notable strengths and limitations. The strengths include; a representative sample of 457 pediatric hospital admissions, with biological samples which were tested for a wide range of bacterial and viral pathogens known to cause pneumonia at a world class research laboratory. The available data on possible predictors included a wide panel of novel and conventional biomarkers (8) along with over 20 clinical signs and symptoms [23]. Despite these strengths, we acknowledge a major limitation is probable outcome misclassification; 54% of the children included in the possible bacterial group in fact had unknown etiology with negative blood culture and negative PCR for all the tested viral targets. This is a direct result of the insensitivity of bacterial cultures and the required specificity of PCR [6,37]. Although bacterial culture is the gold standard for identifying bacterial infections, it is insensitive due to contamination, prior antibiotic therapy and when an infection exists in inaccessible sites [27,37,38]. We performed sensitivity analyses excluding the unknowns, the multivariable model, after controlling for age and chest-wall indrawing also included cough, crackles, and AGT as significant predictors of etiology (Supplementary table 7). However, these factors would not be useful for clinical decision making in ‘real life’, given that there will always be a lot of infections of unknown etiology. Further, the PCR testing panel that was used to define viral pneumonia identified 11 respiratory pathogens. However, there are other viruses known to cause hospitalization with pneumonia such as human bocavirus which were not on the PCR testing panel.

An additional limitation of this analysis is that the laboratory biomarker levels were recorded as MFI units. Although internally valid, this limits the study’s replicability and generalizability and direct comparability to other studies that report their levels and biomarker thresholds as concentrations. Lastly, although our study cohort is representative of pediatric hospital admissions, a large amount of antimicrobial use occurs in outpatient settings and our findings cannot be generalised to outpatient care.

## Conclusion

This study demonstrated that distinguishing between bacterial and viral cases of childhood pneumonia in sub-Saharan Africa remains a challenge. Following the widespread administration of the PCV and Hib vaccines, the case mix on presentation to hospital is increasingly of viral etiology and therefore would not benefit from antibiotics. However, despite having a cohort of over 400 children, and combining clinical presentations with novel and traditional biomarkers in a predictive model, we still could not identify a group of predictors that can accurately identify bacterial infections with enough sensitivity and specificity to guide clinical decision making. These findings highlight the need for diagnostic strategies that explicitly account for disease severity and age-related variation which affect the performance of some biomarkers.

## Supplementary Data

Supplementary materials are available in a separate file consisting of data provided by the authors to benefit the reader and data will be provided upon request to the author or the data governance office at the KWTRP. The posted materials are not copyedited and are the sole responsibility of the authors, so questions or comments should be addressed to the corresponding author.

## Notes

## Data Availability

All data produced in the present study are available upon reasonable request to the authors

## Acknowledgements

The authors appreciate the clinical staff at Kilifi County Referral Hospital and the parents, guardians, and children who participated in this study. This work is published with permission from director KEMRI.

## Author statement

The authors appreciate the clinical staff at Kilifi County Referral Hospital and the parents, guardians, and children who participated in this study. They declare no conflict of interest and that none of the authors or respective institutions received payments or services in the past 36 months from a third party that could be perceived to influence, or give the appearance of potentially influencing, the submitted work. This work was funded by IDeAL and no author or institution at any time received payment or services from a third party for any aspect of the submitted work.

## Author contributions

K.G. designed and conceived the project and supervised the study along with C.J.S. C.M. performed data analysis and wrote the initial draft. E.T.G processed the samples at the KWTRP laboratory. J.M.W and C.J.S conducted the biomarker discovery and validation study which this study builds upon. K.G., C.M., E.T.G., J.M.W., and C.J.S. reviewed the manuscript and produced the final draft.

