## Supplementary tables 1 to 8 and supplementary figure 1 to 4 for "Predictive Modelling to Differentiate Bacterial and Viral cases of Childhood Pneumonia in Kilifi, Kenya using Protein Markers and Clinical Data"

Supplementary Table 1: Continuous variables descriptive statistics at baseline

| Characteristic | Overall  N = 457^1^ | Probable viral  N = 228^1^ | Possible bacterial  N = 229^1^ | p-value^2^ |
| --- | --- | --- | --- | --- |
| **DEMOGRAPHICS** | **Median (IQR)** | **Median (IQR)** | **Median (IQR)** |  |
| Age, months | 9 (5, 19) | 8 (4, 16) | 12 (5, 22) | 0.007 |
| **ANTHROPOMETRIC MESURES** |  |  |  |  |
| Weight | 7.20 (5.90, 9.04) | 7.04 (5.92, 8.78) | 7.44 (5.74, 9.50) | 0.4 |
| Missing | 3 | 1 | 2 |  |
| Length | 69 (62, 78) | 68 (62, 77) | 70 (63, 80) | 0.070 |
| Missing | 21 | 14 | 7 |  |
| Mid-upper arm circumference | 13.00 (12.00, 14.00) | 13.40 (12.00, 14.00) | 13.00 (11.50, 14.00) | 0.002 |
| Missing | 11 | 7 | 4 |  |
| Head circumference | 44.0 (42.0, 47.0) | 44.0 (41.6, 46.6) | 44.8 (42.0, 47.0) | 0.081 |
| Missing | 12 | 8 | 4 |  |
| **CLINICAL SIGNS AND SYMPTOMS** | | | | |
| Heart rate | 163 (146, 180) | 165 (148, 181) | 160 (142, 178) | 0.075 |
| Respiratory rate | 53 (44, 61) | 55 (44, 62) | 51 (43, 60) | 0.2 |
| Missing | 4 | 4 | 0 |  |
| Oxygen saturation | 97.0 (93.0, 99.0) | 96.0 (92.5, 98.0) | 98.0 (94.0, 99.0) | <0.001 |
| Axillary temperature | 38.00 (37.20, 39.00) | 38.10 (37.40, 39.00) | 37.90 (37.00, 38.90) | 0.3 |
| **LABORATORY PARAMETERS** | | | | |
| Hemoglobin (g/dL) | 9.30 (8.10, 10.50) | 9.50 (8.40, 10.50) | 9.00 (7.80, 10.50) | 0.029 |
| Missing | 16 | 9 | 7 |  |
| Mean corpuscular volume (fL) | 69 (62, 75) | 69 (62, 74) | 69 (62, 76) | 0.2 |
| Missing | 18 | 9 | 9 |  |
| Platelets, x𝟏𝟎^3^ /μL | 434 (305, 567) | 434 (322, 549) | 444 (285, 597) | 0.9 |
| Missing | 17 | 9 | 8 |  |
| White Blood Cell count, x𝟏𝟎^3^ /μL | 14 (10, 21) | 13 (10, 18) | 16 (10, 24) | 0.002 |
| Missing | 16 | 9 | 7 |  |
| Red Blood Cell count, x𝟏𝟎^6^/μL | 4.38 (3.83, 4.86) | 4.44 (4.10, 4.89) | 4.25 (3.63, 4.80) | 0.014 |
| Missing | 17 | 9 | 8 |  |
| Neutrophils | 6.0 (3.3, 10.6) | 5.6 (3.3, 8.2) | 6.9 (3.4, 13.2) | 0.017 |
| Missing | 111 | 45 | 66 |  |
| Lymphocytes | 4.3 (2.8, 6.9) | 4.4 (3.0, 7.2) | 4.2 (2.7, 6.4) | 0.2 |
| Missing | 101 | 36 | 65 |  |
| Monocytes | 1.27 (0.77, 2.26) | 1.24 (0.79, 1.95) | 1.30 (0.71, 2.73) | 0.4 |
| Missing | 111 | 45 | 66 |  |
| Eosinophils | 0.13 (0.07, 0.24) | 0.10 (0.06, 0.18) | 0.14 (0.08, 0.29) | <0.001 |
| Missing | 112 | 45 | 67 |  |
| Basophils | 0.21 (0.10, 0.39) | 0.21 (0.10, 0.35) | 0.20 (0.11, 0.44) | 0.6 |
| Missing | 111 | 45 | 66 |  |
| **LABORATORY MARKERS** | | | | |
| AGT | 7.77 (7.52, 8.12) | 7.71 (7.49, 8.02) | 7.86 (7.55, 8.25) | 0.002 |
| CRP | 9.58 (9.34, 9.80) | 9.56 (9.29, 9.77) | 9.60 (9.36, 9.84) | 0.066 |
| HRG | 7.62 (7.31, 7.89) | 7.64 (7.34, 7.93) | 7.60 (7.28, 7.84) | 0.2 |
| LBP | 7.28 (7.05, 7.53) | 7.22 (7.04, 7.47) | 7.34 (7.08, 7.65) | 0.005 |
| PCT | 5.83 (5.59, 6.22) | 5.78 (5.56, 6.15) | 5.86 (5.64, 6.26) | 0.020 |
| PON1 | 6.05 (5.87, 6.32) | 6.03 (5.86, 6.32) | 6.05 (5.88, 6.33) | 0.4 |
| SERPINA1 | 8.54 (8.26, 8.88) | 8.50 (8.23, 8.77) | 8.63 (8.31, 9.00) | 0.002 |
| SERPINA3 | 8.78 (8.47, 9.08) | 8.83 (8.50, 9.14) | 8.71 (8.45, 9.03) | 0.014 |

^The p-values represent the tests of association between the variables and pneumonia etiology, parametric and non-parametric tests of median differences between variables in the bacterial and viral groups. Associations or differences less than 0.05 are considered statistically significant. The Wilcoxon rank sum test was used to test for the differences in the median levels.^

Supplementary Table 2: Categorical variables descriptive statistics at baseline

| Characteristic | Overall  N = 457^1^ | Probable viral  N = 228^1^ | Possible bacterial  N = 229^1^ | p-value^2^ |
| --- | --- | --- | --- | --- |
| **DEMOGRAPHICS** | **N (%)** | **N (%)** | **N (%)** |  |
| Sex |  |  |  | 0.2^c^ |
| Male | 261 (57%) | 123 (54%) | 138 (60%) |  |
| Female | 196 (43%) | 105 (46%) | 91 (40%) |  |
| Age groups |  |  |  | 0.020^c^ |
| 2 - 5 | 143 (31%) | 85 (37%) | 58 (25%) |  |
| 6 - 11 | 116 (25%) | 60 (26%) | 56 (24%) |  |
| 12 - 23 | 108 (24%) | 49 (21%) | 59 (26%) |  |
| 24 - 35 | 51 (11%) | 20 (8.8%) | 31 (14%) |  |
| >= 36 | 39 (8.5%) | 14 (6.1%) | 25 (11%) |  |
| Age groups (Below and above 2yrs) |  |  |  | 0.010^c^ |
| < 24 | 367 (80%) | 194 (85%) | 173 (76%) |  |
| >= 24 | 90 (20%) | 34 (15%) | 56 (24%) |  |
| **CLINICAL SIGNS AND SYMPTOMS** | | | | |
| Pneumonia severity^1^ |  |  |  | <0.001^c^ |
| Pneumonia | 170 (37%) | 64 (28%) | 106 (46%) |  |
| Severe pneumonia | 287 (63%) | 164 (72%) | 123 (54%) |  |
| Muac/weight for age^2^ |  |  |  | 0.014^c^ |
| Well nourished | 254 (76%) | 124 (83%) | 130 (70%) |  |
| Moderate malnutrition | 29 (8.7%) | 7 (4.7%) | 22 (12%) |  |
| Severe malnutrition | 51 (15%) | 18 (12%) | 33 (18%) |  |
| Missing | 123 | 79 | 44 |  |
| Anemia |  |  |  | 0.014^c^ |
| Normal level | 228 (52%) | 128 (58%) | 100 (45%) |  |
| Moderate anemia | 180 (41%) | 79 (36%) | 101 (45%) |  |
| Severe anemia | 33 (7.5%) | 12 (5.5%) | 21 (9.5%) |  |
| Missing | 16 | 9 | 7 |  |
| Presence of oedema |  |  |  | <0.001^f^ |
| No Oedema | 445 (98%) | 228 (100%) | 217 (95%) |  |
| Oedema present | 11 (2.4%) | 0 (0%) | 11 (4.8%) |  |
| Missing | 1 | 0 | 1 |  |
| Elevated heart rate^3^ |  |  |  | 0.7^c^ |
| Normal rate | 166 (36%) | 81 (36%) | 85 (37%) |  |
| Elevated heart rate | 291 (64%) | 147 (64%) | 144 (63%) |  |
| Respiratory distress (tachypnea)^4^ |  |  |  | 0.7^c^ |
| Normal | 120 (26%) | 62 (27%) | 58 (25%) |  |
| Respiratory distress | 337 (74%) | 166 (73%) | 171 (75%) |  |
| Hypoxemia |  |  |  | 0.3^c^ |
| Normal (92–100%) | 377 (82%) | 184 (81%) | 193 (84%) |  |
| Mild hypoxemia (85–91%) | 46 (10%) | 28 (12%) | 18 (7.9%) |  |
| Severe hypoxemia (< 85%) | 34 (7.4%) | 16 (7.0%) | 18 (7.9%) |  |
| Hypoxemia binary |  |  |  | 0.3^c^ |
| No hypoxemia (>= 92%) | 377 (82%) | 184 (81%) | 193 (84%) |  |
| Hypoxemia (< 92%) | 80 (18%) | 44 (19%) | 36 (16%) |  |
| Chest wall indrawing |  |  |  | <0.001^c^ |
| No | 79 (17%) | 10 (4.4%) | 69 (30%) |  |
| Yes | 377 (83%) | 218 (96%) | 159 (70%) |  |
| Missing | 1 | 0 | 1 |  |
| Cough |  |  |  | <0.001^c^ |
| No | 68 (15%) | 9 (3.9%) | 59 (26%) |  |
| Yes | 389 (85%) | 219 (96%) | 170 (74%) |  |
| Diarrhea |  |  |  | 0.011^c^ |
| No | 385 (84%) | 202 (89%) | 183 (80%) |  |
| Yes | 72 (16%) | 26 (11%) | 46 (20%) |  |
| Vomiting |  |  |  | 0.4^c^ |
| No | 367 (80%) | 187 (82%) | 180 (79%) |  |
| Yes | 90 (20%) | 41 (18%) | 49 (21%) |  |
| Convulsion |  |  |  | 0.002^c^ |
| No | 414 (91%) | 216 (95%) | 198 (86%) |  |
| Yes | 43 (9.4%) | 12 (5.3%) | 31 (14%) |  |
| Capillary refill (seconds) |  |  |  | 0.013^c^ |
| >=2 | 223 (49%) | 98 (43%) | 125 (55%) |  |
| <2 | 234 (51%) | 130 (57%) | 104 (45%) |  |
| Fever |  |  |  | 0.2^c^ |
| Axillary temp (<38℃) | 217 (47%) | 102 (45%) | 115 (50%) |  |
| Axillary temp (≥38℃) | 240 (53%) | 126 (55%) | 114 (50%) |  |
| Conscious level |  |  |  | 0.041^c^ |
| Alert | 361 (79%) | 189 (83%) | 172 (75%) |  |
| Impaired | 96 (21%) | 39 (17%) | 57 (25%) |  |
| Pallor |  |  |  | 0.011^c^ |
| No | 362 (79%) | 192 (84%) | 170 (75%) |  |
| Yes | 94 (21%) | 36 (16%) | 58 (25%) |  |
| Missing | 1 | 0 | 1 |  |
| Cyanosis |  |  |  | >0.9^f^ |
| No | 455 (99.8%) | 228 (100%) | 227 (99.6%) |  |
| Yes | 1 (0.2%) | 0 (0%) | 1 (0.4%) |  |
| Missing | 1 | 0 | 1 |  |
| Clubbing |  |  |  | >0.9^f^ |
| No | 454 (99.8%) | 227 (99.6%) | 227 (100%) |  |
| Yes | 1 (0.2%) | 1 (0.4%) | 0 (0%) |  |
| Missing | 2 | 0 | 2 |  |
| Lymphadenopathy |  |  |  | 0.033^f^ |
| No | 441 (98%) | 224 (99%) | 217 (96%) |  |
| Yes | 11 (2.4%) | 2 (0.9%) | 9 (4.0%) |  |
| Missing | 5 | 2 | 3 |  |
| Sunken eye |  |  |  | <0.001^c^ |
| No | 424 (93%) | 221 (97%) | 203 (89%) |  |
| Yes | 32 (7.0%) | 7 (3.1%) | 25 (11%) |  |
| Missing | 1 | 0 | 1 |  |
| Nasal flaring |  |  |  | <0.001^c^ |
| No | 216 (47%) | 89 (39%) | 127 (56%) |  |
| Yes | 240 (53%) | 139 (61%) | 101 (44%) |  |
| Missing | 1 | 0 | 1 |  |
| Stridor |  |  |  | >0.9^c^ |
| No | 444 (97%) | 222 (97%) | 222 (97%) |  |
| Yes | 12 (2.6%) | 6 (2.6%) | 6 (2.6%) |  |
| Missing | 1 | 0 | 1 |  |
| Dull percussion |  |  |  | 0.6^f^ |
| No | 452 (99%) | 227 (100%) | 225 (99%) |  |
| Yes | 4 (0.9%) | 1 (0.4%) | 3 (1.3%) |  |
| Missing | 1 | 0 | 1 |  |
| Crackles |  |  |  | <0.001^c^ |
| No | 444 (97%) | 222 (97%) | 222 (97%) |  |
| Yes | 12 (2.6%) | 6 (2.6%) | 6 (2.6%) |  |
| Missing | 2 | 1 | 1 |  |
| Head nodding |  |  |  | 0.008^c^ |
| No | 389 (87%) | 185 (83%) | 204 (91%) |  |
| Yes | 59 (13%) | 39 (17%) | 20 (8.9%) |  |
| Missing | 9 | 4 | 5 |  |
| Unable to drink/feed |  |  |  | 0.086^c^ |
| No | 416 (93%) | 202 (91%) | 214 (95%) |  |
| Yes | 31 (6.9%) | 20 (9.0%) | 11 (4.9%) |  |
| Missing | 10 | 6 | 4 |  |
| Oral candida/thrush |  |  |  | 0.004^f^ |
| No | 447 (98%) | 228 (100%) | 219 (96%) |  |
| Yes | 9 (2.0%) | 0 (0%) | 9 (3.9%) |  |
| Missing | 1 | 0 | 1 |  |
| Weak pulse volume |  |  |  | 0.2^f^ |
| No | 448 (98%) | 226 (99%) | 222 (97%) |  |
| Yes | 9 (2.0%) | 2 (0.9%) | 7 (3.1%) |  |
| Presence of shock |  |  |  | 0.2^c^ |
| No | 432 (95%) | 219 (96%) | 213 (93%) |  |
| Yes | 25 (5.5%) | 9 (3.9%) | 16 (7.0%) |  |
| Season enrolled |  |  |  | 0.086^c^ |
| April-June | 86 (19%) | 36 (16%) | 50 (22%) |  |
| January-March | 102 (22%) | 54 (24%) | 48 (21%) |  |
| July-September | 148 (32%) | 68 (30%) | 80 (35%) |  |
| October-December | 121 (26%) | 70 (31%) | 51 (22%) |  |
| **LABORATORY PARAMETERS** | | | | |
| Hemoglobin factored^5^ |  |  |  | 0.018^f^ |
| Normal count | 352 (80%) | 184 (84%) | 168 (76%) |  |
| Low levels | 82 (19%) | 30 (14%) | 52 (23%) |  |
| Elevated levels | 7 (1.6%) | 5 (2.3%) | 2 (0.9%) |  |
| Missing | 16 | 9 | 7 |  |
| Mean corpuscular volume (fL) factored^6^ |  |  |  | 0.017^c^ |
| Normal count | 402 (92%) | 199 (91%) | 203 (92%) |  |
| Low levels | 18 (4.1%) | 14 (6.4%) | 4 (1.8%) |  |
| Elevated levels | 19 (4.3%) | 6 (2.7%) | 13 (5.9%) |  |
| Missing | 18 | 9 | 9 |  |
| Platelets factored^7^ |  |  |  | <0.001^c^ |
| Normal count | 227 (52%) | 134 (61%) | 93 (42%) |  |
| Low levels | 20 (4.5%) | 5 (2.3%) | 15 (6.8%) |  |
| Elevated levels | 193 (44%) | 80 (37%) | 113 (51%) |  |
| Missing | 17 | 9 | 8 |  |
| White Blood Cell count factored^8^ |  |  |  | <0.001^c^ |
| Normal count | 231 (52%) | 135 (62%) | 96 (43%) |  |
| Low levels | 22 (5.0%) | 7 (3.2%) | 15 (6.8%) |  |
| Elevated levels | 188 (43%) | 77 (35%) | 111 (50%) |  |
| Missing | 16 | 9 | 7 |  |
| Red Blood Cell count factored^9^ |  |  |  | <0.001^f^ |
| Normal count | 351 (80%) | 191 (87%) | 160 (72%) |  |
| Low levels | 79 (18%) | 25 (11%) | 54 (24%) |  |
| Elevated levels | 10 (2.3%) | 3 (1.4%) | 7 (3.2%) |  |
| Missing | 17 | 9 | 8 |  |
| Neutrophils factored^10^ |  |  |  | 0.2^f^ |
| Normal count | 116 (34%) | 67 (37%) | 49 (30%) |  |
| Elevated levels | 220 (64%) | 113 (62%) | 107 (66%) |  |
| Low levels | 10 (2.9%) | 3 (1.6%) | 7 (4.3%) |  |
| Missing | 111 | 45 | 66 |  |
| Lymphocytes factored^11^ |  |  |  | 0.4^c^ |
| Normal count | 230 (65%) | 128 (67%) | 102 (62%) |  |
| Elevated levels | 36 (10%) | 21 (11%) | 15 (9.1%) |  |
| Low levels | 90 (25%) | 43 (22%) | 47 (29%) |  |
| Missing | 101 | 36 | 65 |  |
| Monocytes factored^12^ |  |  |  | 0.076^f^ |
| Normal count | 212 (61%) | 121 (66%) | 91 (56%) |  |
| Low levels | 5 (1.4%) | 1 (0.5%) | 4 (2.5%) |  |
| Elevated levels | 129 (37%) | 61 (33%) | 68 (42%) |  |
| Missing | 111 | 45 | 66 |  |
| Eosinophils factored^13^ |  |  |  | 0.6^c^ |
| Normal count | 169 (49%) | 93 (51%) | 76 (47%) |  |
| Low levels | 164 (48%) | 85 (46%) | 79 (49%) |  |
| Elevated levels | 12 (3.5%) | 5 (2.7%) | 7 (4.3%) |  |
| Missing | 112 | 45 | 67 |  |
| **LABORATORY MARKERS** | | | | |
| AGT categorical |  |  |  | <0.001^c^ |
| Below (< 7.9144) | 287 (63%) | 164 (72%) | 123 (54%) |  |
| Above (>= 7.9144) | 170 (37%) | 64 (28%) | 106 (46%) |  |
| CRP categorical |  |  |  | 0.13^c^ |
| Below (< 9.6889) | 285 (62%) | 150 (66%) | 135 (59%) |  |
| Above (>= 9.6889) | 172 (38%) | 78 (34%) | 94 (41%) |  |
| HRG categorical |  |  |  | 0.052^c^ |
| Below (< 7.8058) | 312 (68%) | 146 (64%) | 166 (72%) |  |
| Above (>= 7.8058) | 145 (32%) | 82 (36%) | 63 (28%) |  |
| LBP categorical |  |  |  | 0.003^c^ |
| Below (< 7.4234) | 294 (64%) | 162 (71%) | 132 (58%) |  |
| Above (>= 7.4234) | 163 (36%) | 66 (29%) | 97 (42%) |  |
| PCT categorical |  |  |  | 0.003^c^ |
| Below (< 5.8337) | 229 (50%) | 130 (57%) | 99 (43%) |  |
| Above (>= 5.8337) | 228 (50%) | 98 (43%) | 130 (57%) |  |
| PON1 categorical |  |  |  | 0.13^c^ |
| Below (< 5.9306) | 153 (33%) | 84 (37%) | 69 (30%) |  |
| Above (>= 5.9306) | 304 (67%) | 144 (63%) | 160 (70%) |  |
| SERPINA3 categorical |  |  |  | 0.003^c^ |
| Below (< 8.7369) | 210 (46%) | 89 (39%) | 121 (53%) |  |
| Above (>= 8.7369) | 247 (54%) | 139 (61%) | 108 (47%) |  |
| SERPINA1 categorical |  |  |  | <0.001^c^ |
| Below (< 8.6896) | 285 (62%) | 161 (71%) | 124 (54%) |  |
| Above (>= 8.6896) | 172 (38%) | 67 (29%) | 105 (46%) |  |
| ^c^Pearson's Chi-squared test; ^f^Fisher's exact test | | | | |

^The p-values represent the tests of association between the variables and pneumonia etiology, parametric and non-parametric tests of association between variables in the bacterial and viral groups. Associations or differences less than 0.05 are considered statistically significant. Pearson's Chi-squared test and Fisher's exact test were used to test for associations. Fisher’s was applied to variables with a cell count of less than 5.^

^1Non-severe pneumonia = fast breathing or lower chest wall indrawing only. Severe pneumonia = fast breathing or lower chest wall indrawing PLUS at least one danger sign (danger signs are any of the following: cyanosis, head nodding, inability to drink/feed, convulsions, lethargy/ unresponsiveness or impaired consciousness, grunting, nasal flaring, or hypoxemia). Hypoxemia is defined by oxygen saturation less than 92%. 2Muac/weight for age is defined differently for children below and above 6 months of age. For children younger than 6 months, a weight for age z-score of less than -3 indicates severe malnutrition and a weight for age above 2 but less than or equal to -3 indicates moderate malnutrition. For children 6 months of age with a mid-upper circumference less than or equal to 11 indicates severe malnutrition while a mid-upper circumference greater than 11 and less than 12 indicates moderate malnutrition. 3Elevated heart rate is defined by more than 160 beats per minute for children less than 12 months of age, more than 150 beats per minute for children 12 to 35 months of age, and more than 140 beats per minute for children 36 to 59 months of age. 4Respiratory distress was classified according to the children’s ages in months following the WHO guidelines. Children less than 2 months of age with 60 or more breaths per minute, children between 2 and 11 months with 50 and more breaths per minute, and children more than 11 months of age with 40 or more breaths per minute are considered to be having respiratory distress. Hemoglobin, mean corpuscular volume, platelets, neutrophils, platelets, lymphocytes, monocytes, eosinophils are factored according to the reference categorization study carried out in children in Kilifi. The WBC counts are represented on a 95% confidence interval scale with normal counts ranging from (5.14, 14.90) for children less than 5 months of age, (5.97, 17.23) for children between 6 and 11 months of age, and (5.12, 16.28) for children 12 to 59 months of age. WBC levels below the lower limit represent the low levels, while values above the upper limit represent elevated WBC levels. The Red Blood cell count is represented on a 95% confidence interval scale with normal counts ranging from (5.14, 14.90) for children less than 5 months of age, (2.85, 5.34) for children between 6 and 11 months of age, and (3.80, 5.78) for children 12 to 59 months of age. RBC levels below the lower limit represent low levels.^

^[Reference for the categorisation of WBC and RBC: (Downs, L. O., Orindi, B., Hamaluba, M., Bejon, P., Ochola-Oyier, L. I., & Ngetsa, C. (2024). Establishing laboratory reference ranges for adults and children in Kilifi, Kenya. medRxiv, 2024-10.)^

Supplementary Table 3: Continuous variables crude analysis results (Training dataset)

| Characteristic | Overall  N = 319^1^ | Probable viral  N = 159^1^ | Possible bacterial  N = 160^1^ | Crude RR [95% CI] | LRT p-value |
| --- | --- | --- | --- | --- | --- |
| Weight | 7.21 (5.78, 9.15) | 6.99 (5.84, 8.82) | 7.36 (5.70, 9.56) | 0.97 [0.92, 1.02] | 0.2158 |
| Length | 69 (62, 78) | 67 (62, 77) | 71 (61, 81) | 0.99 [0.97, 1.01] | 0.2339 |
| **Mid-upper arm circumference** | **13.00 (12.00, 14.20)** | **13.40 (12.00, 14.30)** | **13.00 (11.40, 14.00)** | **0.96 [0.92, 1.01]** | **0.0842** |
| Head circumference | 44.1 (41.6, 47.0) | 44.0 (41.4, 47.0) | 45.0 (41.9, 47.0) | 0.99 [0.96, 1.02] | 0.4503 |
| Heart rate | 163 (146, 178) | 164 (150, 179) | 162 (141, 178) | 1.00 [1.00, 1.00] | 0.8693 |
| Respiratory rate | 52 (44, 60) | 54 (44, 61) | 50 (42, 60) | 1.00 [0.99, 1.01] | 0.9039 |
| Oxygen saturation | 96 (93, 99) | 95 (92, 98) | 98 (94, 100) | 1.00 [0.99, 1.02] | 0.5605 |
| Axillary temperature | 38.00 (37.20, 39.00) | 38.10 (37.40, 38.90) | 37.85 (36.90, 39.10) | 0.98 [0.91, 1.06] | 0.6980 |
| **LABORATORY PARAMETERS** | | | | | |
| Hemoglobin (g/dL) | 9.40 (8.20, 10.50) | 9.50 (8.60, 10.50) | 9.10 (7.80, 10.50) | 0.99 [0.94, 1.04] | 0.5785 |
| Mean corpuscular volume (fL) | 69 (63, 75) | 69 (63, 74) | 69 (63, 76) | 1.00 [0.99, 1.02] | 0.5099 |
| Platelets, x𝟏𝟎^3^ /μL | 434 (302, 559) | 438 (327, 549) | 432 (247, 559) | 1.00 [1.00, 1.00] | 0.9322 |
| White Blood Cell count, x𝟏𝟎^3^ /μL | 14 (10, 20) | 13 (10, 18) | 16 (10, 24) | 1.00 [1.00, 1.01] | 0.4646 |
| Red Blood Cell count, x𝟏𝟎^6^/μL | 4.35 (3.84, 4.77) | 4.39 (4.11, 4.85) | 4.29 (3.66, 4.75) | 0.97 [0.88, 1.07] | 0.5368 |
| Neutrophils | 6.0 (3.2, 10.7) | 5.3 (3.1, 8.4) | 7.7 (3.6, 13.4) | 1.01 [0.99, 1.02] | 0.2512 |
| Lymphocytes | 4.4 (2.8, 6.9) | 4.5 (3.1, 7.3) | 4.2 (2.6, 6.4) | 1.00 [0.99, 1.01] | 0.9298 |
| Monocytes | 1.27 (0.78, 2.28) | 1.28 (0.79, 1.93) | 1.26 (0.71, 2.84) | 1.01 [0.98, 1.05] | 0.4639 |
| Eosinophils | 0.12 (0.07, 0.22) | 0.10 (0.06, 0.18) | 0.14 (0.08, 0.28) | 1.13 [0.87, 1.41] | 0.3474 |
| Basophils | 0.21 (0.11, 0.41) | 0.20 (0.11, 0.36) | 0.21 (0.10, 0.45) | 1.00 [0.91, 1.06] | 0.9986 |
| **LABORATORY MARKERS** | | | | | |
| **AGT** | **7.79 (7.52, 8.15)** | **7.70 (7.50, 8.04)** | **7.91 (7.52, 8.30)** | **1.12 [0.95, 1.32]** | **0.1786** |
| CRP | 9.57 (9.35, 9.79) | 9.56 (9.35, 9.78) | 9.59 (9.36, 9.82) | 1.10 [0.86, 1.42] | 0.4581 |
| HRG | 7.63 (7.32, 7.92) | 7.64 (7.33, 7.95) | 7.61 (7.30, 7.84) | 1.00 [0.82, 1.23] | 0.9618 |
| LBP | 7.28 (7.04, 7.52) | 7.22 (7.03, 7.46) | 7.34 (7.07, 7.68) | 1.10 [0.89, 1.36] | 0.3768 |
| PCT | 5.83 (5.56, 6.22) | 5.76 (5.53, 6.16) | 5.86 (5.61, 6.26) | 1.08 [0.90, 1.28] | 0.4061 |
| PON1 | 6.04 (5.87, 6.33) | 6.02 (5.86, 6.32) | 6.06 (5.89, 6.35) | 1.06 [0.83, 1.35] | 0.6502 |
| SERPINA3 | 8.79 (8.48, 9.07) | 8.83 (8.53, 9.13) | 8.71 (8.45, 9.04) | 1.00 [0.85, 1.19] | 0.9562 |
| SERPINA1 | 8.54 (8.26, 8.85) | 8.51 (8.24, 8.75) | 8.59 (8.31, 9.01) | 1.11 [0.91, 1.37] | 0.2921 |

^Variables with an LRT p-value <0.2 are further analysed/considered for inclusion in the multi variable model.^

Supplementary Table 4: Categorical variables crude analysis results (Training set)

| Characteristic | Overall  N = 319^1^ | Probable viral  N = 159^1^ | Possible bacterial  N = 160^1^ | Crude RR [95% CI] | LRT p-value |
| --- | --- | --- | --- | --- | --- |
| DEMPOGRAPHIC | | | | | |
| Sex |  |  |  |  | 0.9088 |
| Male | 177 (55%) | 87 (55%) | 90 (56%) | Ref |  |
| Female | 142 (45%) | 72 (45%) | 70 (44%) | 0.99 [0.82, 1.19] |  |
| Age groups |  |  |  |  | 0.7391 |
| 2 - 5 | 98 (31%) | 60 (38%) | 38 (24%) | Ref |  |
| 6 - 11 | 86 (27%) | 44 (28%) | 42 (26%) | 1.07 [0.84, 1.37] |  |
| 12 - 23 | 72 (23%) | 33 (21%) | 39 (24%) | 1.11 [0.86, 1.43] |  |
| 24 - 35 | 35 (11%) | 14 (8.8%) | 21 (13%) | 1.15 [0.84, 1.56] |  |
| >= 36 | 28 (8.8%) | 8 (5.0%) | 20 (13%) | 1.24 [0.88, 1.70] |  |
| Season enrolled |  |  |  |  | 0.8404 |
| April-June | 63 (20%) | 24 (15%) | 39 (24%) | Ref |  |
| January-March | 67 (21%) | 35 (22%) | 32 (20%) | 0.93 [0.70, 1.22] |  |
| July-September | 101 (32%) | 49 (31%) | 52 (33%) | 0.95 [0.74, 1.23] |  |
| October-December | 88 (28%) | 51 (32%) | 37 (23%) | 0.89 [0.68, 1.16] |  |
| Fever |  |  |  |  | 0.5223 |
| Axillary temp (<38℃) | 157 (49%) | 73 (46%) | 84 (53%) | Ref |  |
| Axillary temp (≥38℃) | 162 (51%) | 86 (54%) | 76 (48%) | 0.94 [0.79, 1.13] |  |
| Conscious level |  |  |  |  | 0.4153 |
| Alert | 251 (79%) | 132 (83%) | 119 (74%) | Ref |  |
| Impaired | 68 (21%) | 27 (17%) | 41 (26%) | 1.09 [0.88, 1.35] |  |
| Presence of oedema |  |  |  |  | 0.2947 |
| No Oedema | 310 (97%) | 159 (100%) | 151 (94%) | Ref |  |
| Oedema present | 9 (2.8%) | 0 (0%) | 9 (5.6%) | 1.30 [0.78, 2.04 |  |
| Elevated heart rate^1^ |  |  |  |  | 0.9221 |
| Normal rate | 116 (36%) | 56 (35%) | 60 (38%) | Ref |  |
| Elevated heart rate | 203 (64%) | 103 (65%) | 100 (63%) | 0.99 [0.82, 1.20] |  |
| Respiratory distress (tachypnea)^2^ |  |  |  |  | 0.9553 |
| Normal | 86 (27%) | 44 (28%) | 42 (26%) | Ref |  |
| Respiratory distress | 233 (73%) | 115 (72%) | 118 (74%) | 1.01 [0.82, 1.24] |  |
| Hypoxemia |  |  |  |  | 0.6088 |
| Normal (92–100%) | 267 (84%) | 129 (81%) | 138 (86%) | Ref |  |
| Mild hypoxemia (85–91%) | 27 (8.5%) | 19 (12%) | 8 (5.0%) | 0.85 [0.59, 1.18] |  |
| Severe hypoxemia (< 85%) | 25 (7.8%) | 11 (6.9%) | 14 (8.8%) | 1.04 [0.74, 1.43] |  |
| Hypoxemia binary |  |  |  |  | 0.6330 |
| No hypoxemia (>= 92%) | 267 (84%) | 129 (81%) | 138 (86%) | Ref |  |
| Hypoxemia (< 92%) | 52 (16%) | 30 (19%) | 22 (14%) | 0.94 [0.73, 1.20] |  |
| **Chest wall indrawing** |  |  |  |  | **0.0081** |
| **No** | **65 (20%)** | **7 (4.4%)** | **58 (36%)** | **Ref** |  |
| **Yes** | **254 (80%)** | **152 (96%)** | **102 (64%)** | **0.75 [0.61, 0.93]** |  |
| Pneumonia severity^3^ |  |  |  |  | 0.2121 |
| Pneumonia | 127 (40%) | 48 (30%) | 79 (49%) | Ref |  |
| Severe pneumonia | 192 (60%) | 111 (70%) | 81 (51%) | 0.89 [0.74, 1.07] |  |
| Capillary refill (seconds) |  |  |  |  | 0.4168 |
| >=2 | 160 (50%) | 70 (44%) | 90 (56%) | Ref |  |
| <2 | 159 (50%) | 89 (56%) | 70 (44%) | 0.93 [0.78, 1.11] |  |
| Muac/weight for age^4^ |  |  |  |  | 0.5562 |
| Well nourished | 175 (74%) | 85 (80%) | 90 (69%) | Ref |  |
| Moderate malnutrition | 19 (8.1%) | 5 (4.7%) | 14 (11%) | 1.21 [0.80, 1.79] |  |
| Severe malnutrition | 42 (18%) | 16 (15%) | 26 (20%) | 1.12 [0.84, 1.47] |  |
| Anemia |  |  |  |  | 0.7644 |
| Normal level | 162 (53%) | 88 (58%) | 74 (48%) | Ref |  |
| Moderate anemia | 124 (40%) | 58 (38%) | 66 (43%) | 1.05 [0.86, 1.27] |  |
| Severe anemia | 22 (7.1%) | 7 (4.6%) | 15 (9.7%) | 1.13 [0.78, 1.59] |  |
| **Cough** |  |  |  |  | **0.0143** |
| **No** | **54 (17%)** | **5 (3.1%)** | **49 (31%)** | **Ref** |  |
| **Yes** | **265 (83%)** | **154 (97%)** | **111 (69%)** | **0.75 [0.60, 0.94]** |  |
| Diarrhea |  |  |  |  | 0.2180 |
| No | 277 (87%) | 147 (92%) | 130 (81%) | Ref |  |
| Yes | 42 (13%) | 12 (7.5%) | 30 (19%) | 1.18 [0.91, 1.51] |  |
| Vomiting |  |  |  |  | 0.9124 |
| No | 255 (80%) | 128 (81%) | 127 (79%) | Ref |  |
| Yes | 64 (20%) | 31 (19%) | 33 (21%) | 1.01 [0.80, 1.26] |  |
| **Convulsion** |  |  |  |  | **0.1456** |
| **No** | **285 (89%)** | **153 (96%)** | **132 (83%)** | **Ref** |  |
| **Yes** | **34 (11%)** | **6 (3.8%)** | **28 (18%)** | **1.23 [0.93, 1.59]** |  |
| Pallor |  |  |  |  | 0.6138 |
| No | 251 (79%) | 131 (82%) | 120 (75%) | Ref |  |
| Yes | 68 (21%) | 28 (18%) | 40 (25%) | 1.06 [0.85, 1.31] |  |
| Cyanosis |  |  |  |  | 0.6241 |
| No | 318 (100%) | 159 (100%) | 159 (99%) | Ref |  |
| Yes | 1 (0.3%) | 0 (0%) | 1 (0.6%) | 1.45 [0.24, 4.54] |  |
| Sunken eye |  |  |  |  | 0.2759 |
| No | 295 (92%) | 154 (97%) | 141 (88%) | Ref |  |
| Yes | 24 (7.5%) | 5 (3.1%) | 19 (12%) | 1.20 [0.86, 1.62] |  |
| Drinks thirstily |  |  |  |  | 0.4785 |
| No | 168 (98%) | 86 (100%) | 82 (96%) | Ref |  |
| Yes | 3 (1.8%) | 0 (0%) | 3 (3.5%) | 1.36 [0.53, 2.84] |  |
| Nasal flaring |  |  |  |  | 0.2904 |
| No | 165 (52%) | 69 (43%) | 96 (60%) | Ref |  |
| Yes | 154 (48%) | 90 (57%) | 64 (40%) | 0.91 [0.76, 1.09] |  |
| Stridor |  |  |  |  | 0.7942 |
| No | 312 (98%) | 156 (98%) | 156 (98%) | Ref |  |
| Yes | 7 (2.2%) | 3 (1.9%) | 4 (2.5%) | 1.08 [0.56, 1.88] |  |
| **Crackles** |  |  |  |  | **0.1218** |
| **No** | **202 (63%)** | **83 (52%)** | **119 (74%)** | **Ref** |  |
| **Yes** | **117 (37%)** | **76 (48%)** | **41 (26%)** | **0.86 [0.71, 1.04]** |  |
| Head nodding |  |  |  |  | 0.2667 |
| No | 278 (88%) | 129 (82%) | 149 (94%) | Ref |  |
| Yes | 37 (12%) | 28 (18%) | 9 (5.7%) | 0.84 [0.61, 1.14] |  |
| Unable to drink/feed |  |  |  |  | 0.6335 |
| No | 294 (94%) | 142 (92%) | 152 (96%) | Ref |  |
| Yes | 20 (6.4%) | 13 (8.4%) | 7 (4.4%) | 0.91 [0.60, 1.32] |  |
| Oral candida/thrush |  |  |  |  | 0.2186 |
| No | 312 (98%) | 159 (100%) | 153 (96%) | Ref |  |
| Yes | 7 (2.2%) | 0 (0%) | 7 (4.4%) | 1.43 [0.79, 2.35] |  |
| Presence of shock |  |  |  |  | 0.6102 |
| No | 298 (93%) | 151 (95%) | 147 (92%) | Ref |  |
| Yes | 21 (6.6%) | 8 (5.0%) | 13 (8.1%) | 1.10 [0.76, 1.53] |  |
| Weak pulse volume |  |  |  |  | 0.6114 |
| No | 312 (98%) | 157 (99%) | 155 (97%) | Ref |  |
| Yes | 7 (2.2%) | 2 (1.3%) | 5 (3.1%) | 1.17 [0.62, 1.98] |  |
| **LABORATORY PARAMTERS** | | | | | |
| Hemoglobin factored^5^ |  |  |  |  | 0.5654 |
| Normal count | 250 (81%) | 132 (86%) | 118 (76%) | Ref |  |
| Low levels | 52 (17%) | 17 (11%) | 35 (23%) | 1.12 [0.87, 1.41] |  |
| Elevated levels | 6 (1.9%) | 4 (2.6%) | 2 (1.3%) | 0.84 [0.38, 1.61] |  |
| Mean corpuscular volume (fL)^6^ factored |  |  |  |  | 0.6475 |
| Normal count | 282 (92%) | 140 (92%) | 142 (93%) | Ref |  |
| Low levels | 9 (2.9%) | 8 (5.2%) | 1 (0.7%) | 0.76 [0.34, 1.35] |  |
| Elevated levels | 15 (4.9%) | 5 (3.3%) | 10 (6.5%) | 1.06 [0.68, 1.56] |  |
| Platelets factored^7^ |  |  |  |  | 0.5026 |
| Normal count | 161 (52%) | 96 (63%) | 65 (42%) | Ref |  |
| Low levels | 15 (4.9%) | 3 (2.0%) | 12 (7.8%) | 1.36 [0.80, 2.20] |  |
| Elevated levels | 131 (43%) | 54 (35%) | 77 (50%) | 1.22 [0.77, 1.84] |  |
| White Blood Cell count factored^8^ |  |  |  |  | 0.3808 |
| Normal count | 161 (52%) | 94 (61%) | 67 (43%) | Ref |  |
| Low levels | 17 (5.5%) | 5 (3.3%) | 12 (7.7%) | 1.22 [0.81, 1.77] |  |
| Elevated levels | 130 (42%) | 54 (35%) | 76 (49%) | 1.12 [0.93, 1.35] |  |
| Red Blood Cell count factored^9^ |  |  |  |  | 0.4744 |
| Normal count | 245 (80%) | 135 (88%) | 110 (71%) | Ref |  |
| Low levels | 54 (18%) | 16 (10%) | 38 (25%) | 1.14 [0.89, 1.45] |  |
| Elevated levels | 8 (2.6%) | 2 (1.3%) | 6 (3.9%) | 1.20 [0.67, 1.99] |  |
| Neutrophils factored^10^ |  |  |  |  | 0.5701 |
| Normal count | 79 (32%) | 51 (40%) | 28 (24%) | Ref |  |
| Elevated levels | 158 (65%) | 76 (59%) | 82 (71%) | 1.14 [0.89, 1.41] |  |
| Low levels | 7 (2.9%) | 2 (1.6%) | 5 (4.3%) | 1.26 [0.66, 2.21] |  |
| Lymphocytes factored^11^ |  |  |  |  | 0.8248 |
| Normal count | 159 (63%) | 90 (67%) | 69 (59%) | Ref |  |
| Elevated levels | 25 (10.0%) | 14 (10%) | 11 (9.5%) | 1.02 [0.70, 1.44] |  |
| Low levels | 67 (27%) | 31 (23%) | 36 (31%) | 1.08 [0.85, 1.35] |  |
| Monocytes factored^12^ |  |  |  |  | 0.8211 |
| Normal count | 152 (62%) | 86 (67%) | 66 (57%) | Ref |  |
| Low levels | 4 (1.6%) | 1 (0.8%) | 3 (2.6%) | 1.25 [0.53, 2.45] |  |
| Elevated levels | 88 (36%) | 42 (33%) | 46 (40%) | 1.04 [0.83, 1.29] |  |
| Eosinophils factored^13^ |  |  |  |  | 0.6805 |
| Normal count | 122 (50%) | 67 (52%) | 55 (48%) | Ref |  |
| Low levels | 113 (47%) | 59 (46%) | 54 (47%) | 0.88 [0.64, 1.20] |  |
| Elevated levels | 8 (3.3%) | 3 (2.3%) | 5 (4.4%) | 1.03 [0.55, 1.77] |  |
| **LABORATORY MARKERS** | | | | | |
| **AGT categorical** |  |  |  |  | **0.1119** |
| **Below (< 7.9144)** | **195 (61%)** | **114 (72%)** | **81 (51%)** | **Ref** |  |
| **Above (>= 7.9144)** | **124 (39%)** | **45 (28%)** | **79 (49%)** | **1.16 [0.97, 1.39]** |  |
| CRP categorical |  |  |  |  | 0.6935 |
| Below (< 9.6889) | 197 (62%) | 103 (65%) | 94 (59%) | Ref |  |
| Above (>= 9.6889) | 122 (38%) | 56 (35%) | 66 (41%) | 1.04 [0.86, 1.25] |  |
| HRG categorical |  |  |  |  | 0.621 |
| Below (< 7.8058) | 215 (67%) | 101 (64%) | 114 (71%) | Ref |  |
| Above (>= 7.8058) | 104 (33%) | 58 (36%) | 46 (29%) | 0.95 [0.78, 1.15] |  |
| LBP categorical |  |  |  |  | 0.3711 |
| Below (< 7.4234) | 206 (65%) | 114 (72%) | 92 (58%) | Ref |  |
| Above (>= 7.4234) | 113 (35%) | 45 (28%) | 68 (43%) | 1.09 [0.90, 1.31] |  |
| PCT categorical |  |  |  |  | 0.4788 |
| Below (< 5.8337) | 163 (51%) | 90 (57%) | 73 (46%) | Ref |  |
| Above (>= 5.8337) | 156 (49%) | 69 (43%) | 87 (54%) | 1.07 [0.89, 1.28] |  |
| PON1 categorical |  |  |  |  | 0.4794 |
| Below (< 5.9306) | 107 (34%) | 61 (38%) | 46 (29%) | Ref |  |
| Above (>= 5.9306) | 212 (66%) | 98 (62%) | 114 (71%) | 1.07 [0.89, 1.30] |  |
| SERPINA3 categorical |  |  |  |  | 0.3452 |
| Below (< 8.7369) | 143 (45%) | 60 (38%) | 83 (52%) | Ref |  |
| Above (>= 8.7369) | 176 (55%) | 99 (62%) | 77 (48%) | 0.92 [0.77, 1.10] |  |
| SERPINA1 categorical |  |  |  |  | 0.2853 |
| Below (< 8.6896) | 201 (63%) | 112 (70%) | 89 (56%) | Ref |  |
| Above (>= 8.6896) | 118 (37%) | 47 (30%) | 71 (44%) | 1.11 [0.92, 1.33] |  |
| **AGT (at one year cut-off)** |  |  |  |  | **0.1077** |
| **Low** | **182 (57%)** | **107 (67%)** | **75 (47%)** | **Ref** |  |
| **High** | **137 (43%)** | **52 (33%)** | **85 (53%)** | **1.16 [0.97, 1.39]** |  |
| CRP (at one year cut-off) |  |  |  |  | 0.6226 |
| Low | 204 (64%) | 108 (68%) | 96 (60%) | Ref |  |
| High | 115 (36%) | 51 (32%) | 64 (40%) | 1.05 [0.87, 1.26] |  |
| PCT (at one year cut-off) |  |  |  |  | 0.4596 |
| Low | 167 (52%) | 92 (58%) | 75 (47%) | Ref |  |
| High | 152 (48%) | 67 (42%) | 85 (53%) | 1.07 [0.89, 1.28] |  |
| LBP (at one year cut-off) |  |  |  |  | 0.3137 |
| Low | 226 (71%) | 123 (77%) | 103 (64%) | Ref |  |
| High | 93 (29%) | 36 (23%) | 57 (36%) | 1.11 [0.91, 1.34] |  |
| HRG (at one year cut-off) |  |  |  |  | 0.5569 |
| Low | 178 (56%) | 79 (50%) | 99 (62%) | Ref |  |
| High | 141 (44%) | 80 (50%) | 61 (38%) | 0.95 [0.78, 1.14] |  |
| PON1 (at one year cut-off) |  |  |  |  | 0.3775 |
| Low | 171 (54%) | 99 (62%) | 72 (45%) | Ref |  |
| High | 148 (46%) | 60 (38%) | 88 (55%) | 1.09 [0.90, 1.32] |  |
| **SERPINA1 (at one year cut-off)** |  |  |  |  | **0.1053** |
| **Low** | **227 (71%)** | **129 (81%)** | **98 (61%)** | **Ref** |  |
| **High** | **92 (29%)** | **30 (19%)** | **62 (39%)** | **1.20 [0.94, 1.39]** |  |
| SERPINA3 (at one year cut-off) |  |  |  |  | 0.3082 |
| Low | 179 (56%) | 79 (50%) | 100 (63%) | Ref |  |
| High | 140 (44%) | 80 (50%) | 60 (38%) | 1.10 [0.92, 1.32] |  |
| **AGT (at two years cut-off)** |  |  |  |  | **0.1514** |
| **Low** | **166 (52%)** | **97 (61%)** | **69 (43%)** | **Ref** |  |
| **High** | **153 (48%)** | **62 (39%)** | **91 (57%)** | **1.14 [0.95, 1.37]** |  |
| CRP (at two years cut-off) |  |  |  |  | 0.5243 |
| Low | 210 (66%) | 114 (72%) | 96 (60%) | Ref |  |
| High | 109 (34%) | 45 (28%) | 64 (40%) | 1.07 [0.88, 1.29] |  |
| PCT (at two years cut-off) |  |  |  |  | 0.5572 |
| Low | 187 (59%) | 102 (64%) | 85 (53%) | Ref |  |
| High | 132 (41%) | 57 (36%) | 75 (47%) | 1.06 [0.88, 1.27] |  |
| LBP (at two years cut-off) |  |  |  |  | 0.3428 |
| Low | 209 (66%) | 116 (73%) | 93 (58%) | Ref |  |
| High | 110 (34%) | 43 (27%) | 67 (42%) | 1.10 [0.91, 1.32] |  |
| HRG (at two years cut-off) |  |  |  |  | 0.8855 |
| Low | 86 (27%) | 41 (26%) | 45 (28%) | Ref |  |
| High | 233 (73%) | 118 (74%) | 115 (72%) | 0.99 [0.81, 1.21] |  |
| PON1 (at two years cut-off) |  |  |  |  | 0.3559 |
| Low | 126 (39%) | 70 (44%) | 56 (35%) | Ref |  |
| High | 193 (61%) | 89 (56%) | 104 (65%) | 1.09 [0.91, 1.32] |  |
| SERPINA1 (at two years cut-off) |  |  |  |  | 0.2600 |
| Low | 202 (63%) | 70 (44%) | 56 (35%) | Ref |  |
| High | 117 (37%) | 89 (56%) | 104 (65%) | 1.11 [0.92, 1.33] |  |
| SERPINA3 (at two years cut-off) |  |  |  |  | 0.5542 |
| Low | 240 (75%) | 113 (71%) | 127 (79%) | Ref |  |
| High | 79 (25%) | 46 (29%) | 33 (21%) | 0.94 [0.75, 1.16] |  |
| **AGT (at three years cut-off)** |  |  |  |  | **0.1581** |
| **Low** | **164 (51%)** | **96 (60%)** | **68 (43%)** | **Ref** |  |
| **High** | **155 (49%)** | **63 (40%)** | **92 (58%)** | **1.14 [0.95, 1.36]** |  |
| CRP (at three years cut-off) |  |  |  |  | 0.6939 |
| Low | 190 (60%) | 101 (64%) | 89 (56%) | Ref |  |
| High | 129 (40%) | 58 (36%) | 71 (44%) | 1.04 [0.86, 1.25] |  |
| PCT (at three years cut-off) |  |  |  |  | 0.4459 |
| Low | 161 (50%) | 90 (57%) | 71 (44%) | Ref |  |
| High | 158 (50%) | 69 (43%) | 89 (56%) | 1.07 [0.90, 1.28] |  |
| LBP (at three years cut-off) |  |  |  |  | 0.4867 |
| Low | 211 (66%) | 116 (73%) | 95 (59%) | Ref |  |
| High | 108 (34%) | 43 (27%) | 65 (41%) | 1.07 [0.88, 1.30] |  |
| HRG (at three years cut-off) |  |  |  |  | 0.9350 |
| Low | 87 (27%) | 42 (26%) | 45 (28%) | Ref |  |
| High | 232 (73%) | 117 (74%) | 115 (72%) | 0.99 [0.81, 1.22] |  |
| PON1 (at three years cut-off) |  |  |  |  | 0.3929 |
| Low | 112 (35%) | 64 (40%) | 48 (30%) | Ref |  |
| High | 207 (65%) | 95 (60%) | 112 (70%) | 1.09 [0.90, 1.32] |  |
| SERPINA1 (at three years cut-off) |  |  |  |  | 0.2743 |
| Low | 200 (63%) | 112 (70%) | 88 (55%) | Ref |  |
| High | 119 (37%) | 47 (30%) | 72 (45%) | 1.11 [0.92, 1.33] |  |
| SERPINA3 (at three years cut-off) |  |  |  |  | 0.4968 |
| Low | 150 (47%) | 65 (41%) | 85 (53%) | Ref |  |
| High | 169 (53%) | 94 (59%) | 75 (47%) | 0.94 [0.78, 1.13] |  |

^Variables with an LRT p-value of <0.2 were carried forward to be assessed in the multivariate model. All crude RRs were adjusted for age as an a-priori variable.^ ^1Elevated heart rate is defined by more than 160 beats per minute for children less than 12 months of age, more than 150 beats per minute for children 12 to 35 months of age, and more than 140 beats per minute for children 36 to 59 months of age. 2Respiratory distress was classified according to the children’s ages in months following the WHO guidelines. Children less than 2 months of age with 60 or more breaths per minute, children between 2 and 11 months with 50 and more breaths per minute, and children more than 11 months of age with 40 or more breaths per minute are considered to be having respiratory distress. 3Non-severe pneumonia = fast breathing or lower chest wall indrawing only. Severe pneumonia = fast breathing or lower chest wall indrawing PLUS at least one danger sign (danger signs are any of the following: cyanosis, head nodding, inability to drink/feed, convulsions, lethargy/ unresponsiveness or impaired consciousness, grunting, nasal flaring, or hypoxemia). Hypoxemia is defined by oxygen saturation less than 92%. 4Muac/weight for age is defined differently for children below and above 6 months of age. For children younger than 6 months, a weight for age z-score of less than -3 indicates severe malnutrition and a weight for age above 2 but less than or equal to -3 indicates moderate malnutrition. For children 6 months of age with a mid-upper circumference less than or equal to 11 indicates severe malnutrition while a mid-upper circumference greater than 11 and less than 12 indicates moderate malnutrition. Hemoglobin, mean corpuscular volume, platelets, neutrophils, platelets, lymphocytes, monocytes, eosinophils are factored according to the reference categorization study carried out in children in Kilifi. The WBC counts are represented on a 95% confidence interval scale with normal counts ranging from (5.14, 14.90) for children less than 5 months of age, (5.97, 17.23) for children between 6 and 11 months of age, and (5.12, 16.28) for children 12 to 59 months of age. WBC levels below the lower limit represent the low levels, while values above the upper limit represent elevated WBC levels. The Red Blood cell count is represented on a 95% confidence interval scale with normal counts ranging from (5.14, 14.90) for children less than 5 months of age, (2.85, 5.34) for children between 6 and 11 months of age, and (3.80, 5.78) for children 12 to 59 months of age. RBC levels below the lower limit represent low levels.^

^[Reference for the categorisation of WBC and RBC: (Downs, L. O., Orindi, B., Hamaluba, M., Bejon, P., Ochola-Oyier, L. I., & Ngetsa, C. (2024). Establishing laboratory reference ranges for adults and children in Kilifi, Kenya. medRxiv, 2024-10.)^

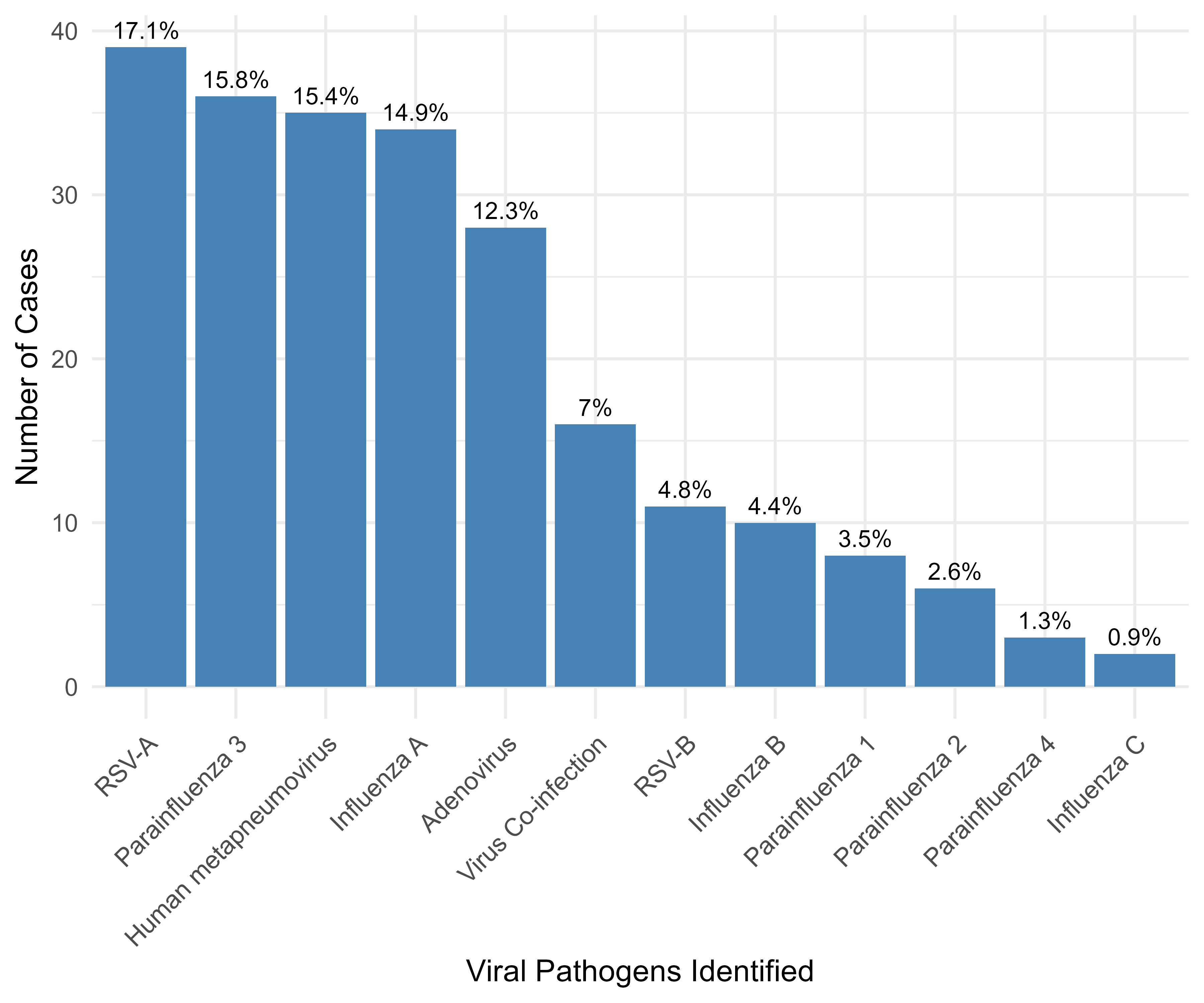

Supplementary Figure 1: Viral pathogens identified in the entire study population

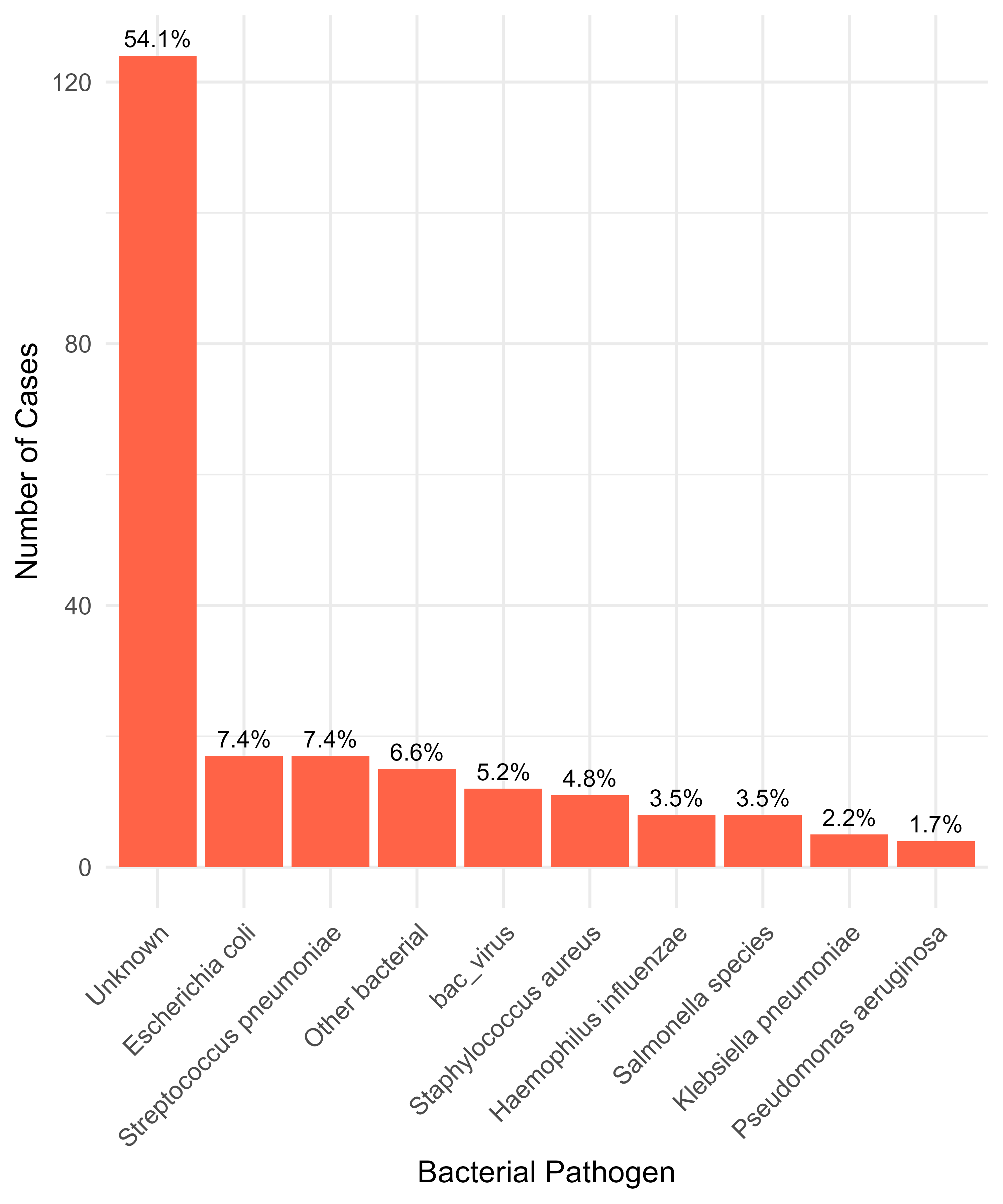

Supplementary Figure 2: Bacterial pathogens identified in the entire study population

54% of the unknown infections were classified as bacterial cases. These are pathogens that tested negative on both blood culture and PCR. Further, they include viral pathogens such as rhinovirus and HCOV strains (OC43, NL63, 229E) that present like pneumonia but do not truly cause pneumonia.

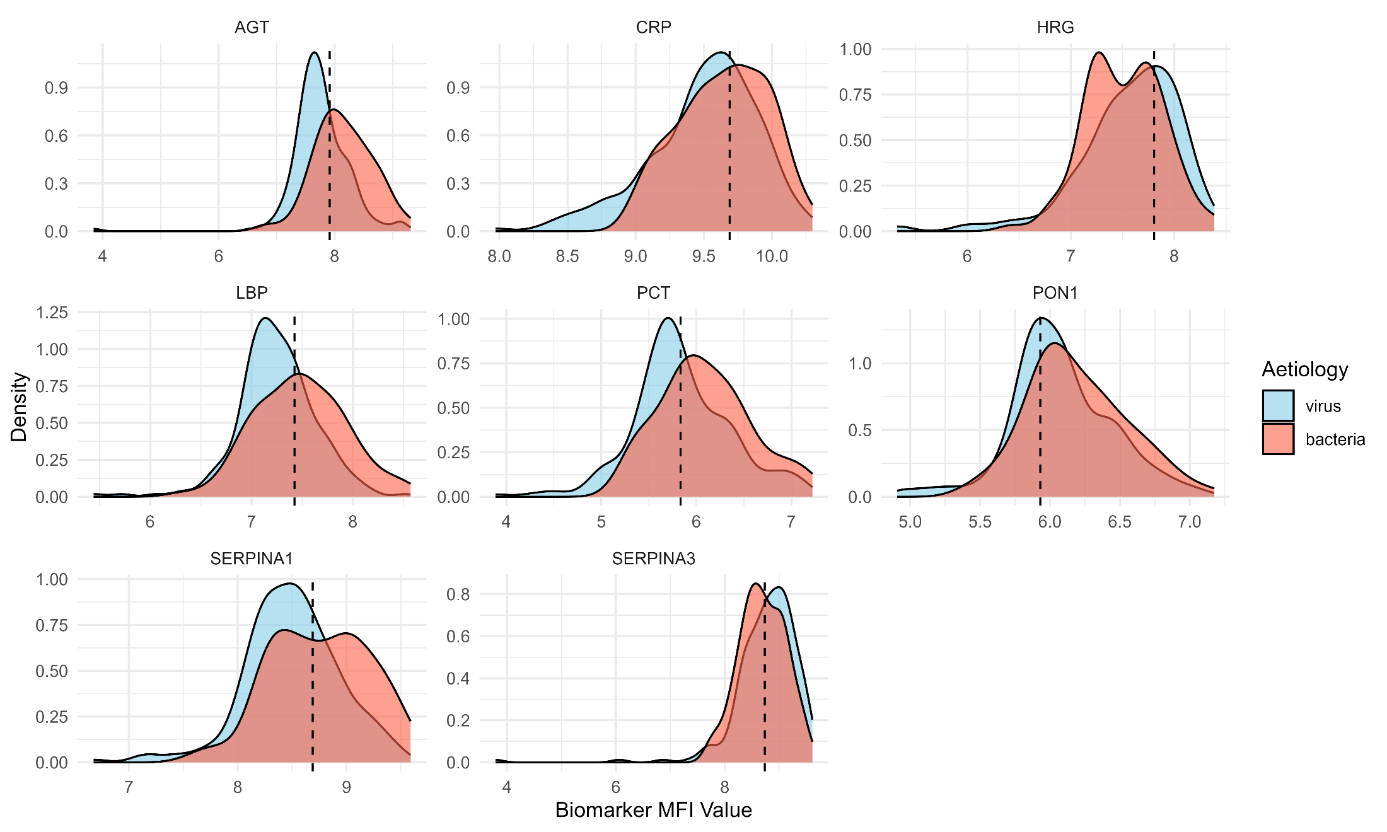

Supplementary Figure 3: Density plot restricted to only bacterial and viral infections

The above density plot indicates the biomarker MFI levels of each biomarker with respective thresholds that separate bacterial from viral infections. However, we observe a significant degree of overlap between the biomarker levels of the bacterial and viral infections leading the thresholds to not reliably distinguish between bacterial and viral infections with enough sensitivity and specificity.

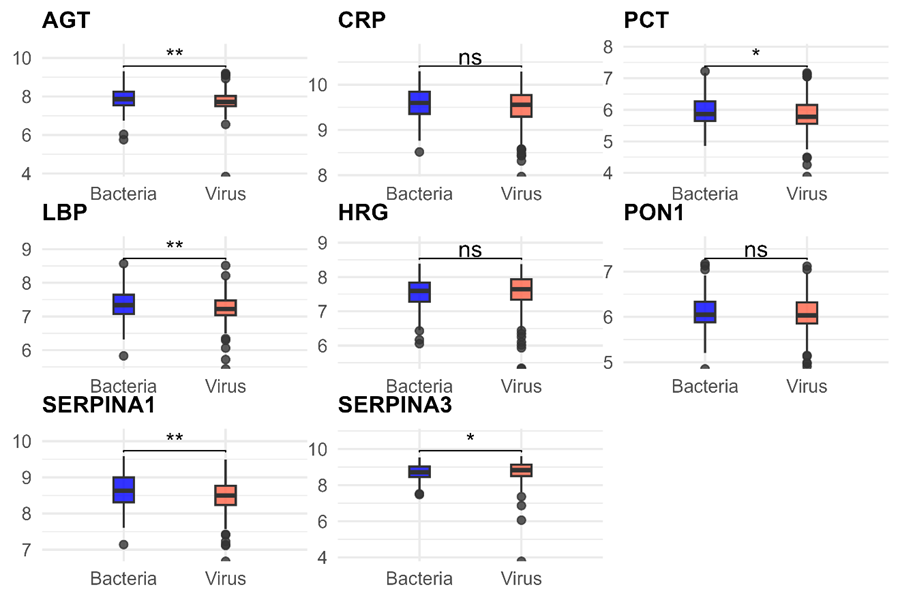

Supplementary Figure 4: Boxplots of median differences in the biomarker MFI values

CRP, HRG, and PON1 have insignificant differences in their median MFI values while AGT, PCT, LBP, SERPINA1, and SERPINA3 have significant differences in their median MFI values. AGT, LBP, PCT, and SERPINA1 have higher MFI median values in bacterial infections compared to viral infections while SERPINA3 has higher median MFI value in viral infections compared to bacterial infections.

Supplementary Table 5: Biomarker thresholds and performance in pneumonia etiology differentiation in bacterial and viral pneumonia infections (excluding the unknowns).

| **Biomarker** | **Threshold (MFI)** | **Sensitivity** | **Specificity** | **PPV** | **NPV** | **AUC [95% CI]** |
| --- | --- | --- | --- | --- | --- | --- |
| AGT | 7.9144 | 0.67 | 0.72 | 0.49 | 0.84 | 0.73 [0.67, 0.80] |
| CRP | 9.6889 | 0.52 | 0.66 | 0.38 | 0.77 | 0.60 [0.53, 0.67] |
| PCT | 5.8337 | 0.72 | 0.57 | 0.41 | 0.83 | 0.64 [0.57, 0.71] |
| LBP | 7.4234 | 0.57 | 0.71 | 0.45 | 0.80 | 0.65 [0.58, 0.72] |
| HRG | 7.8058 | 0.78 | 0.36 | 0.33 | 0.80 | 0.57 [0.50, 0.64] |
| PON1 | 5.9306 | 0.80 | 0.37 | 0.34 | 0.82 | 0.59 [0.52, 0.66] |
| SERPINA1 | 8.6896 | 0.56 | 0.71 | 0.44 | 0.80 | 0.66 [0.59, 0.73] |
| SERPINA3 | 8.7369 | 0.56 | 0.61 | 0.37 | 0.77 | 0.58 [0.51, 0.65] |
|  | **Threshold (counts)** | **Sensitivity** | **Specificity** | **PPV** | **NPV** | **AUC [95% CI]** |
| WBC | 23.55 | 0.38 | 0.88 | 0.57 | 0.78 | 0.58 [0.50, 0.66] |
| Neutrophils | 11.01 | 0.44 | 0.85 | 0.51 | 0.81 | 0.59 [0.49, 0.68] |
| Lymphocytes | 3.88 | 0.64 | 0.42 | 0.27 | 0.77 | 0.50 [0.41, 0.58] |

In a sub-analysis restricted to only bacterial and viral cases of pneumonia, AGT had moderate discriminatory power (AUC = 0.73, 95% CI [0.67, 0.80]). None of the other novel biomarkers could adequately distinguish between confirmed bacterial and viral cases: SERPINA1 (AUC = 0.66, 95% CI [0.59, 0.73]), LBP (AUC = 0.65, 95% CI [0.58, 0.72]), PCT (AUC = 0.64, 95% CI [0.58, 0.71]), and CRP (AUC = 0.60, 95% CI [0.53, 0.67].

Supplementary Table 6: Final model results on the validation dataset

| Matuli et al. Model (Modified Poisson Regression model) | | | | |
| --- | --- | --- | --- | --- |
| Predictor | **aRR [95% CI]** |  |  |  |
| Age group |  |  |  |  |
| 2 - 5 | Ref |  |  |  |
| 6 - 11  12 - 23 | 1.02 [0.69, 1.49] |  |  |  |
|  | 1.06 [0.73, 1.53] |  |  |  |
| 24 - 35 | 1.09 [0.67, 1.72] |  |  |  |
| ≥ 36 | 1.00 [0.56, 1.69] |  |  |  |
| Chest-wall indrawing |  |  |  |  |
| No | Ref |  |  |  |
| Yes | 0.84 [0.55, 1.33] |  |  |  |
| FARIDA ET. AL MODEL (Multivariate Logistic Regression Model) | | | | |
| Using Farida’s cut-off | | Using our study cut-off | | |
| Predictor | **aRR [95% CI]** |  |  |  |
| CRP |  | **CRP** |  |  |
| Below (< 5.70mg/L) | Ref | **Below (< 9.6889 MFI)** | Ref |  |
| Above (≥ 5.70mg/L) | 1.72 [0.59, 5.24] | **Above (>= 9.6889 MFI)** | 1.38 [0.94, 2.03] |  |
| Fever (≥ 38℃) |  | **Fever (>= 38℃)** |  |  |
| No | Ref | **No** | Ref |  |
| Yes | 0.62 [0.23, 1.58] | **Yes** | 0.78 [0.54, 1.13] |  |
| Season |  | **Season** |  |  |
| Dry season | Ref | **Dry season** | Ref |  |
| Rainy season | 1.11 [0.42, 2.93] | **Rainy season** | 0.82 [0.56, 1.20] |  |
| BHUIYAN ET AL. MODEL (Modified Poisson Regression model) | | | | |
| Using Bhuiyan’s cut-off | | Using our study cut-off | | |
| Predictor | **aRR [95% CI]** |  |  |  |
| CRP |  | **CRP** |  |  |
| Below (<72.0mg/L) | Ref | **Below (< 9.6889 MFI)** | Ref |  |
| Above (≥ 72.0mg/L) | 1.32 [0.92, 1.89] | **Above (>= 9.6889 MFI)** | 1.16 [0.96, 1.38] |  |
| Fever (>= 38℃) |  | **Fever (>= 38℃)** |  |  |
| No | Ref | **No** | Ref |  |
| Yes | 0.92 [0.64, 1.31] | **Yes** | 0.89 [0.74, 1.02] |  |
| MODEL VALIDATION SPECIFICATIONS | | | | |
| Model | **Predictors** | **Sensitivity** | **Specificity** | **AUC [95% CI]** |
| Testing set | Age, chest wall indrawing | 0.50 | 0.68 | 0.61 [0.52, 0.70] |
| Farida et al. model | CRP, fever, rainy season | 0.55 | 0.63 | 0.58 [0.45, 0.71] |
| Bhuiyan et al. model | CRP, fever | 0.55 | 0.86 | 0.74 [0.64, 0.85] |

^The model validation specifications are based on each study’s case definition. For instance, the metrics obtained for the Farida et al. study were obtained when the model was validated on our data that had CRP levels converted to concentrations. The same dataset was applied in validating the Bhuiyan et al. study.^

The Farida et al. study was conducted in Indonesian children 2 – 59 months of age. Their final logistic regression model included fever, CRP (5.70mg/L) and rainy season. When validated on our data, the model could not discriminate between bacterial and viral infections beyond random chance with an AUC of 0.58. The model’s calibration intercept was -1.42 pointing to possible overestimation with a calibration slope of 3.85 indicating possible model underfit.

The Bhuiyan et al. study was carried out in Western Australian children ≤17 years of age. The study recommended the combination of elevated CRP levels with clinical presentations to improve the discrimination between bacterial and viral cases of pneumonia. This study utilized Bhuiyan’s CRP cutoff of ≥ 72mg/L with fever (≥ 38̊C) in a modified Poisson regression model to determine how best these predictors perform in our setting. This model moderately discriminated (AUC = 0.74, 95% CI [0.64, 0.85]) between bacterial and viral pneumonia with a sensitivity of 0.55 and a specificity of 0.86. The model attained a calibration intercept of approximately zero indicating that the model was not miscalibrated and a calibration slope of approximately 1 indicating that the predicted risks align well the observed outcomes.

Supplementary Table 7: Predictors of pneumonia etiology in the multivariable modified Poisson model excluding the unknowns.

| Characteristics | Crude RR [95% CI] | LRT p-value | Adjusted RR [95% CI] | LRT p-value |
| --- | --- | --- | --- | --- |
| Weight | 0.96 [0.91, 1.02] | 0.1952 |  |  |
| Mid upper arm circumference | 0.95 [0.90, 1.00] | 0.0393 |  |  |
| Hemoglobin (g/dL) | 0.96 [0.92, 1.01] | 0.1122 |  |  |
| Red Blood Cell count, x𝟏𝟎^6^/μL | 0.91 [0.82, 1.00] | 0.0585 |  |  |
| Neutrophils | 1.01 [1.00, 1.02] | 0.1753 |  |  |
| Monocytes | 1.03 [0.99, 1.06] | 0.1235 |  |  |
| AGT | 1.27 [1.05, 1.53] | 0.0121 |  |  |
| LBP | 1.22 [0.97, 1.55] | 0.0889 |  |  |
| PCT | 1.15 [0.96, 1.38] | 0.1396 |  |  |
| SERPINA1 | 1.21 [0.98, 1.50] | 0.0810 |  |  |
| Age groups |  | 1.0000 |  |  |
| 2 - 5 | Ref |  |  |  |
| 6 - 11 | 1.00 [0.74, 1.36] |  |  |  |
| 12 - 23 | 1.13 [0.84, 1.51] |  |  |  |
| 24 - 35 | 1.02 [0.62, 1.60] |  |  |  |
| >= 36 | 1.17 [0.73, 1.79] |  |  |  |
| Chest wall indrawing |  | <0.0001 |  | 0.0149 |
| No | Ref |  | Ref |  |
| Yes | 0.61 [0.48, 0.75] |  | 0.70 [0.53, 0.93] |  |
| Pneumonia severity |  | 0.0350 |  |  |
| Pneumonia | Ref |  |  |  |
| Severe pneumonia | 0.81 [0.66, 0.98] |  |  |  |
| Cough |  | 0.0004 |  | 0.0180 |
| No | Ref |  | Ref |  |
| Yes | 0.65 [0.52, 0.82] |  | 0.73 [0.57, 0.95] |  |
| Diarrhea |  | 0.1877 |  |  |
| No | Ref |  |  |  |
| Yes | 1.19 [0.92, 1.51] |  |  |  |
| Convulsion |  | 0.0599 |  |  |
| No | Ref |  |  |  |
| Yes | 1.32 [0.99, 1.74] |  |  |  |
| Pallor |  | 0.1324 |  |  |
| No | Ref |  |  |  |
| Yes | 1.19 [0.95, 1.49] |  |  |  |
| Sunken eye |  | 0.1386 |  |  |
| No | Ref |  |  |  |
| Yes | 1.31 [0.91, 1.81] |  |  |  |
| Nasal flaring |  | 0.0319 |  |  |
| No | Ref |  |  |  |
| Yes | 0.81 [0.66, 0.98] |  |  |  |
| Crackles |  | 0.0185 |  | 0.0202 |
| No | Ref |  | Ref |  |
| Yes | 0.78 [0.63, 0.96] |  | 0.78 [0.63, 0.96] |  |
| Platelets factored |  | 0.1819 |  |  |
| Normal count | Ref |  |  |  |
| Low levels | 1.64 [0.96, 2.65] |  |  |  |
| Elevated levels | 1.34 [0.86, 2.01] |  |  |  |
| White Blood Cell count factored |  | 0.0811 |  |  |
| Normal count | Ref |  |  |  |
| Low levels | 1.46 [1.98, 2.10] |  |  |  |
| Elevated levels | 1.18 [0.96, 1.45] |  |  |  |
| Red Blood Cell count factored |  | 0.0937 |  |  |
| Normal count | Ref |  |  |  |
| Low levels | 1.31 [1.02, 1.66] |  |  |  |
| Elevated levels | 1.22 [0.57, 2.23] |  |  |  |
| AGT categorical |  | 0.0115 |  | 0.0132 |
| Below (< 7.9144) | Ref |  | Ref |  |
| Above (>= 7.9144) | 1.29 [1.06, 1.56] |  | 1.28 [1.05, 1.56] |  |
| LBP categorical |  | 0.0909 |  |  |
| Below (< 7.4234) | Ref |  |  |  |
| Above (>= 7.4234) | 1.19 [0.97, 1.45] |  |  |  |
| PCT categorical |  | 0.0758 |  |  |
| Below (< 5.8337) | Ref |  |  |  |
| Above (>= 5.8337) | 1.19 [0.98, 1.45] |  |  |  |
| SERPINA1 categorical |  | 0.0958 |  |  |
| Below (< 8.6896) | Ref |  |  |  |
| Above (>= 8.6896) | 1.18 [0.97, 1.44] |  |  |  |
| AGT (at one year cut-off) |  | 0.0079 |  |  |
| Low | Ref |  |  |  |
| High | 1.30 [1.07, 1.58] |  |  |  |
| PCT (at one year cut-off) |  | 0.0643 |  |  |
| Low | Ref |  |  |  |
| High | 1.20 [0.99, 1.46] |  |  |  |
| LBP (at one year cut-off) |  | 0.0558 |  |  |
| Low | Ref |  |  |  |
| High | 1.22 [1.00, 1.49] |  |  |  |
| SERPINA1 (at one year cut-off) |  | 0.0704 |  |  |
| Low | Ref |  |  |  |
| High | 1.21 [0.98, 1.49] |  |  |  |
| AGT (at two years cut-off) |  | 0.0115 |  |  |
| Low | Ref |  |  |  |
| High | 1.28 [1.06, 1.56] |  |  |  |
| PCT (at two years cut-off) |  | 0.0695 |  |  |
| Low | Ref |  |  |  |
| High | 1.20 [0.99, 1.46] |  |  |  |
| LBP (at two years cut-off) |  | 0.0687 |  |  |
| Low | Ref |  |  |  |
| High | 1.20 [0.99, 1.47] |  |  |  |
| PON1 (at two years cut-off) |  | 0.1790 |  |  |
| Low | Ref |  |  |  |
| High | 1.15 [0.94, 1.43] |  |  |  |
| SERPINA1 (at two years cut-off) |  | 0.0900 |  |  |
| Low | Ref |  |  |  |
| High | 1.19 [0.97, 1.44] |  |  |  |
| AGT (at three years cut-off) |  | 0.0124 |  |  |
| Low | Ref |  |  |  |
| High | 1.28 [1.05, 1.56] |  |  |  |
| PCT (at three years cut-off) |  | 0.0667 |  |  |
| Low | Ref |  |  |  |
| High | 1.20 [0.99, 1.46] |  |  |  |
| LBP (at three years cut-off) |  | 0.1257 |  |  |
| Low | Ref |  |  |  |
| High | 1.18 [0.95, 1.45] |  |  |  |
| SERPINA1 (at three years cut-off) |  | 0.0856 |  |  |
| Low | Ref |  |  |  |
| High | 1.19 [0.98, 1.45] |  |  |  |

This analysis was carried out using a sample size of 321 bacterial (93) and viral (228) infections to assess how the inclusion of unknowns as bacterial infections affect our model. The table presents the multivariable model results of significant predictors of pneumonia etiology using only bacterial and viral infections (excluding the unknowns). This data was not split into a training and test sets. All the 321 samples were used to fit the model. The final multivariable model had AGT, chest-wall indrawing, cough, and crackles after adjusting for age as the significant predictors of pneumonia etiology. This model has good discrimination (AUC = 0.85, 95% CI [0.80, 0.91], sensitivity = 0.70, specificity = 0.91]. Factors such as AGT which are represented in different forms, were not all included in the multivariable model. The AGT form, say categorised or continuous, was considered for inclusion in the multivariable model if they had the smallest likelihood ratio test (lrt) p-value in crude analysis.

*Supplementary Table 8: Exploratory analysis of biomarker interactions with pneumonia severity*

| **LABORATORY MARKERS** | **Interaction RR** | **p-value** |
| --- | --- | --- |
| **AGT** | **0.85** | **0.0080** |
| **CRP** | **0.77** | **0.0073** |
| PCT | 0.92 | 0.1897 |
| LBP | 0.97 | 0.6837 |
| PON1 | 0.89 | 0.2462 |
| **HRG** | **0.85** | **0.0380** |
| **SERPINA1** | **0.80** | **0.0058** |
| SERPINA3 | 0.91 | 0.1562 |

These biomarkers are in continuous form with their values as MFIs. The significant interactions indicate that the biomarkers are predictive in non-severe cases.
